# Immune profiling demonstrates a common immune signature of delayed acquired immunodeficiency in patients with various etiologies of severe injury

**DOI:** 10.1101/2021.03.12.21253466

**Authors:** Fabienne Venet, Julien Textoris, Sophie Blein, Mary-Luz Rol, Maxime Bodinier, Bertrand Canard, Pierre Cortez, Boris Meunier, Lionel K Tan, Craig Tipple, Laurence Quemeneur, Frédéric Reynier, Philippe Leissner, Christophe Védrine, Yves Bouffard, Benjamin Delwarde, Olivier Martin, Thibaut Girardot, Cyrille Truc, Andrew D. Griffiths, Virginie Moucadel, Alexandre Pachot, Guillaume Monneret, Thomas Rimmelé, for the REALISM study group

## Abstract

**Background and Research Question:** The host response plays a central role in the pathophysiology of sepsis and severe injuries. So far, no study has comprehensively described the overtime changes of the injury-induced immune profile in a large cohort of critically ill patients with different etiologies.

**Study Design and Methods:** 353 septic, trauma and surgical patients and 175 healthy volunteers were prospectively included in the REAnimation Low Immune Status Marker (REALISM) study. Extensive immune profiling was performed by assessing cellular phenotypes and functions, protein and mRNA levels at days 1-2, 3-4 and 5-7 after inclusion using a panel of 30 standardized immune markers.

**Results:** Using REALISM immunomonitoring panel, no specificity in the immune profile was observed between septic, trauma and surgical patients. This common injury-induced immune response was characterized by an initial adaptive (i.e. physiologic) response engaging all constituents of the immune system (pro- and anti-inflammatory cytokine release, innate and adaptive immune responses) but not associated with increased risk of secondary infections. In contrary, the persistence in a subgroup of patients of profound immune alterations at the end of the first week after admission was associated with increased risk of secondary infections independently of exposure to invasive devices. The combined monitoring of markers of pro/anti-inflammatory, innate and adaptive immune responses allowed a better enrichment of patients with risk of secondary infections in the selected population.

**Interpretation:** These results illustrate the delayed development of a common maladaptive injury-acquired immunodeficiency in a subgroup of severely injured patients independently of initial etiologies. Critically ill patients’ immune status could be captured through the combined monitoring a common panel of complementary markers of pro/anti-inflammatory, innate and adaptive immune responses. Such immune monitoring panel will help clinicians to identify critically ill patients who could benefit from tailored immunoadjuvant therapies.

**Clinical Trial Registration:** clinicaltrials.gov: NCT02638779

**Summary conflict of interest statements:** JT, SB, VM and AP are employees of bioMérieux SA, an *in vitro* diagnostic company. FV, TR, YB, BD, OM, TG, CT and GM are employees of Hospices Civils de Lyon. JT, TR, SB, VM, AP, FV and GM work in a joint research unit, co funded by the Hospices Civils de Lyon and bioMérieux. JT, AP, GM and FV are co-inventors in patent applications covering the following markers: CX3CR1, CD127, IL10 and S100A9. LKT and CT are employees of and hold stock and shares in GlaxoSmithKline. LQU is an employee of Sanofi Pasteur. PC was employee of Sanofi, Inc. and declares no other competing interests.

**Funding information:** This study received funding from the Agence Nationale de la Recherche through a grant awarded to BIOASTER (Grant number #ANR-10-AIRT-03) and from bioMérieux, Sanofi and GSK.

## Introduction

Sepsis, defined as a life-threatening organ dysfunction caused by a dysregulated host response to infection, represents a major healthcare challenge with high incidence, morbidity and mortality^1-3^. A recent epidemiological study estimated that, in 2017, the incidence of sepsis worldwide was close to 50 million cases leading to 11 million deaths^4^. Sepsis has therefore been recently acknowledged as a worldwide health priority by the World Health Organization^5^.

The host immune response plays a central role in sepsis pathophysiology. In particular, following the initial overwhelming pro-inflammatory response (cytokine storm) leading to organ failure and shock, sepsis is secondarily associated with the development of immune dysfunction affecting both innate and adaptive immune responses^6,7^. In addition, in patients who survive the initial hyper-inflammatory phase, reactivation of latent viruses, development of secondary infections with opportunistic pathogens and death are frequently observed^7-9^. Consequently, the use of host-targeted therapies which are aimed at rejuvenating the function of the immune response is currently debated in sepsis^10^. However, in the absence of clear-cut clinical signs indicating the immune status of a patient, there is an unmet need to identify markers available on a routine basis to monitor septic patients’ immune status and identify those who could benefit from tailored immunoadjuvant therapies.

Beyond sepsis, similar immune alterations have been described in other cohorts of critically ill patients such as those with severe trauma, burns or surgery^11-13^. Although the modulation of some specific immune parameters showed similarities with the immune alterations observed in sepsis, the possible specificity of the immune profile induced by different types of sterile *vs.* infectious injuries has never been evaluated.

In this context, the primary objective of the REAnimation Low Immune Status Marker (REALISM) study was to perform an extensive immune profiling of injury-induced immune response in a large cohort of critically ill patients with different etiologies. A panel of 30 standardized markers assessing pro/anti-inflammatory, innate and adaptive immune responses was evaluated. Techniques, available in most routine labs, were selected to reflect complementary aspects of the immune responses (i.e. cell numbers, phenotype, transcriptome and function as well as circulating concentrations of cytokines). Secondary objectives included the evaluation of the association between injury-induced immune alterations and secondary infections as a proxy of immune failure. Collectively, we intended to evaluate whether a common immune signature of injury-induced immune alterations could be defined to help rationalize immune monitoring of ICU patients.

## Methods

Detailed study design and experimental protocols have been published previously.14 However, the key aspects are summarized below.

### Prospective study design

We performed a prospective, observational cohort study of critically ill patients who presented with either sepsis, severe trauma or planned surgery to the Anesthesiology and Intensive Care Medicine department of the Edouard Herriot Hospital (Hospices Civils de Lyon, France), for a 28-month inclusion period (ending March 2018). The study protocol was approved by the IRB (Comité de Protection des Personnes Sud-Est II) under number 2015-42-2. This clinical study was also registered at clinicaltrials.gov (NCT02638779).

Inclusion criteria were: patients aged > 18 years, clinical diagnosis of sepsis as defined by 2016 SEPSIS-3 consensus guidelines^1^, severe trauma with injury severity score (ISS) > 15 or surgical patients undergoing major surgeries such as oesophago-gastrectomy, bladder resection with Brickers’ reconstruction, cephalic pancreaticoduodenectomy and abdominal aortic aneurysm surgery by laparotomy. Exclusion criteria were any of the following: presence of a preexistent condition or treatment that could influence patients’ immune status, pregnancy, institutionalized patients, inability to obtain informed consent.

A cohort of 175 healthy volunteers aged from 18 to 82 years (81 males and 94 females) was also recruited prospectively. To account for the possible influence of age and sex on immune parameters, the distribution of healthy volunteers was based on the age and sex demographic data for the French population in 2016.

Written informed consent was obtained from every healthy volunteer and patient upon inclusion. If a patient was unable to consent directly, informed consent was obtained from the patient’s legally authorized representative and reconfirmed from the patient at the earliest opportunity.

### Sampling schedule and collection

In septic and trauma patients, clinical samples and data were collected three times during the first week after admission: at day 1 or 2 (D1-2), D3 or 4 (D3-4) and D5, D6 or D7 (D5-7). One sample was collected in healthy volunteers during the study visit and clinical data were recorded.

Peripheral whole blood was collected for each patient and healthy volunteer in one EDTA tube, two heparin tubes and one PAXgene blood RNA tube (PreAnalytix, Hilden, Germany) at each time point. Tubes were immediately transferred to the lab and processed within 3h after blood sampling. PAXgene samples were stabilized for at least 2h at room temperature after collection and frozen at -80°C following manufacturer’s recommendations. They were subsequently used for mRNA concentration measurements. The EDTA samples were used for flow cytometry immune phenotyping and plasma cytokine level measurements. Heparinized blood was used for functional tests (proliferation, cytokine production experiments).

### Data collection

Patients’ demographics, comorbidities, diagnosis, severity and clinical outcome were manually curated in a prospective fashion. Longitudinal follow-up was performed for a period of 90 days. The following data were recorded: demographic data (age, gender, BMI), disease severity measured by the Simplified Acute Physiological Score (SAPS) II at ICU admission^15^ and the Sequential Organ Failure Assessment (SOFA) score^16^ measured at D1-2, D3-4 and D5-7. The chronic health status was defined using the Charlson comorbidity index^17^ as well as MacCabe score^18^. ISS was recorded for trauma patients^19^. Site and type of infections were recorded at admission for septic patients. Hospital and ICU lengths of stay (LOS) and survival were recorded after 30 and 90 days as well as follow-up location after ICU discharge.

### Definitions of outcomes and secondary infections

The primary outcome was 30-day secondary infection. During the hospital stay, patients were monitored daily for the use of an invasive medical device (tracheal intubation, indwelling urinary catheter and central venous line) and for secondary infections. Information collected about infections was reviewed by an independent adjudication committee composed of three clinicians not involved in study patients’ recruitment or care. Confirmation of the occurrence of secondary infection by this committee was based on guidelines of the European Society of Clinical Microbiology and Infectious Diseases and the Infectious Diseases Society of America. Only the first episode of secondary infection was considered in the analyses. The adjudication committee remained blinded for results of immune parameters.

### Immune profiling panel

The complete list of immune parameters is presented in e-Table 1 and has been detailed elsewhere.^14^ Briefly, the immune profiling panel included the concomitant assessment of parameters reflecting three complementary aspects of the immune response: magnitude of pro- and anti-inflammatory cytokine production, phenotypic, transcriptional and functional profiles of innate (neutrophils, monocytes) and adaptive (lymphocytes) cellular responses.

**Table 1.** Cohort description by subgroup at inclusion. BMI: Body Mass Index. SAPS II: Simplified Acute Physiological Score II. SOFA: Sequential Organ Failure Assessment score. D: Day. Cat: catecholamine. MV: mechanical ventilation. RRT: Renal Replacement Therapy. ICU: Intensive Care Unit. G: Giga. GCS: Glasgow coma scale. HAI: health care associated infection. Hosp: hospital. Data are presented as numbers and percentages (qualitative variables) and medians and 25th/75th percentiles (quantitative variables). Clinical and biological parameters were compared between subgroups using Kruskal-Wallis test.

| Variable | Whole cohort<br>n=353 | Sepsis<br>n=107 | Trauma<br>n=137 | Surgery<br>n=109 | P-value |
| --- | --- | --- | --- | --- | --- |
| Age (year) | 60 [47-71] | 68 [59-77] | 47 [29-57] | 67 [57-73] | < 0.001 |
| Gender (Female) | 122 (34.6 %) | 38 (35.5 %) | 34 (24.8 %) | 50 (45.9 %) | 0.003 |
| BMI (kg/m<sup>2</sup>) | 25.0 [22.3-28.4] | 25.5 [22.7-30.4] | 25.0 [22.6-28.5] | 24.2 [21.9-27.1] | 0.029 |
| **Clinical scores** |  |  |  |  |  |
| Mac Cabe |  |  |  |  | < 0.001 |
| Non fatal disease | 243 (68.8 %) | 66 (61.7 %) | 135 (98.5 %) | 42 (38.5 %) |  |
| Ultimately fatal disease | 98 (27.8 %) | 34 (31.8 %) | 2 (1.5 %) | 62 (56.9 %) |  |
| Rapidly fatal disease | 12 (3.4 %) | 7 (6.5 %) | 0 (0.0 %) | 5 (4.6 %) |  |
| Charlson score | 1 [0-2] | 2 [1-3] | 0 [0-0] | 2 [2-3] | < 0.001 |
| SAPS II score | 29 [20-43] | 47 [38-55] | 26 [20-37] | 20 [14-26] | < 0.001 |
| SOFA score at D1 | 5 [1-8] | 9 [7-11] | 2 [1-7] | 2 [1-4] | < 0.001 |
| SOFA score at D3-4 | 2 [0-5] | 6 [4-9] | 1 [0-2] | 1 [0-2] | < 0.001 |
| SOFA score at D5-7 | 1 [0-2] | 3 [1-5] | 0 [0-1] | 0 [0-1] | < 0.001 |
| **Organ failures at D1** |  |  |  |  |  |
| Vasopressors | 193 (54.7 %) | 100 (93.5 %) | 63 (46.0 %) | 30 (27.5 %) | < 0.001 |
| D30 Cat. free days | 28 [26-29] | 27 [25-29] | 29 [28-30] | 29 [28-29] | < 0.001 |
| Hydrocortisone | 38 (11 %) | 27 (25 %) | 7 (5 %) | 4 (4 %) | < 0.001 |
| Mechanical ventilation | 164 (46 %) | 76 (71 %) | 70 (51 %) | 18 (17 %) | < 0.001 |
| D30 MV free days | 28 [22-29] | 26 [4-29] | 29 [25-29] | 29 [29-29] | < 0.001 |
| PaO<sub>2</sub>/FiO<sub>2</sub> | 245 [174-343] | 210 [159-292] | 291 [197-400] | 318 [297-390] | < 0.001 |
| Urine output (L/24h) | 1.6 [1.0-2.0] | 1.2 [0.5-2.0] | 1.7 [1.2-2.2] | 1.4 [1.1-2.0] | < 0.001 |
| RRT | 61 (17 %) | 42 (39 %) | 11 (8 %) | 8 (7 %) | < 0.001 |
| D30 RRT free days | 14 [0-28] | 5 [0-26] | 27 [10-29] | 27 [25-28] | 0.008 |
| Coma (GCS < 8) | 26 (7.4 %) | 6 (5.6 %) | 20 (14.6 %) | 0 (0.0 %) | < 0.001 |
| **Lab values at D1** |  |  |  |  |  |
| pH | 7.36 [7.30-7.40] | 7.37 [7.31-7.42] | 7.36 [7.30-7.39] | 7.35 [7.30-7.38] | 0.298 |
| Lactate (mM) | 2.2 [1.7-3.3] | 2.2 [1.7-3.5] | 2.2 [1.5-3.1] | 2.1 [1.8-2.9] | 0.741 |
| Creatinine (μM) | 70 [52-106] | 153 [100-237] | 82 [68-102] | 74 [56-114] | < 0.001 |
| Bilirubin (μM) | 16 [10-35] | 19 [12-43] | 12 [7-17] | 16 [10-27] | 0.022 |
| ASAT (UI/L) | 95 [49-192] | 42 [21-110] | 69 [35-157] | 125 [65-249] | 0.52 |
| Leukocytes (G/L) | 13.2 [10.2-17.3] | 13.3 [8.6-22.0] | 13.8 [10.6-17.2] | 12.6 [10.6-15.4] | 0.513 |
| Hemoglobin (g/L) | 116 [101-132] | 112 [92-126] | 123 [109-138] | 110 [98-125] | < 0.001 |
| Platelets (G/L) | 205 [145-267] | 212 [144-306] | 195 [156-238] | 215 [175-267] | 0.048 |
| Neutrophils (G/L) | 10.9 [8.0-14.8] | 11.6 [7.5-20.3] | 11.3 [8.1-14.4] | 10.4 [8.4-12.8] | 0.181 |
| Lymphocytes (G/L) | 1.2 [0.8-1.8] | 1.0 [0.6-1.5] | 1.3 [0.9-2.0] | 1.2 [0.9-1.7] | < 0.001 |
| Monocytes (G/L) | 0.9 [0.7-1.3] | 0.8 [0.5-1.4] | 1.0 [0.8-1.3] | 1.0 [0.7-1.3] | 0.080 |
| **Outcomes** |  |  |  |  |  |
| At least one HAI | 74 (21.0 %) | 20 (18.7 %) | 23 (16.8 %) | 31 (28.4 %) | 0.065 |
| Median HAI onset day | 9 [6-15] | 12 [9-17] | 8 [6-13] | 8 [6-16] | 0.152 |
| D30 ICU free days | 23 [18-26] | 21 [9-25] | 24 [20-26] | 26 [23-28] | < 0.001 |
| D30 hosp. free days | 15 [2-21] | 0 [0-14] | 16 [8-22] | 17 [8-23] | < 0.001 |
| D30 mortality | 20 (5.7 %) | 18 (16.8 %) | 2 (1.5 %) | 0 (0.0 %) | < 0.001 |
| D90 mortality | 33 (9.4 %) | 27 (25.2 %) | 5 (3.7 %) | 1 (0.9 %) | < 0.001 |
| **D30 follow-up (location)** |  |  |  |  | < 0.001 |
| ICU | 21 (6.3 %) | 8 (8.9 %) | 10 (7.4 %) | 3 (2.8 %) |  |
| Continuing care unit | 7 (2.1 %) | 2 (2.2 %) | 1 (0.7 %) | 4 (3.7 %) |  |
| Short-term care | 79 (23.7 %) | 40 (44.5 %) | 17 (12.6 %) | 22 (20.2 %) |  |
| Long-term care | 65 (19.5 %) | 9 (10.0 %) | 47 (34.8 %) | 9 (8.3 %) |  |
| Home | 162 (48.5 %) | 31 (34.4 %) | 60 (44.5 %) | 71 (65.1 %) |  |

### Statistical analyses

Data are presented as numbers and percentages (qualitative variables) and medians and 25th/75th percentiles (quantitative variables). Chi square or Fisher’s exact test were used for qualitative variables assessment. Quantitative variables were compared with Mann-Whitney *U* test or Student t tests. For each immune parameter, reference intervals were calculated based on 2.5th and 97.5th percentiles of healthy volunteers’ values.

For all pairs of immune parameters measured at D1-2, Spearman’s rho correlation coefficients were estimated and summarized in a correlation matrix.

Differences between markers and subgroups were estimated using ANalysis of COVAriance (ANCOVA) models at each time point. They were adjusted on age, SOFA score at admission and Charlson score and p-values were corrected for multi-testing using the Bonferroni method. PCA based on all makers at each time-point were also performed to estimate the overlap between subgroups. Patients with missing value for any markers were excluded. Mean silhouette score over the two first PCA axes was reported for each subgroup at each time point, as a surrogate of subgroups overlap.

The associations between D30 secondary infection and immune parameters (with death as competitive risk) were performed by implementing univariate Fine and Gray models. These models were applied only at D1-2 due to the insufficient number of deaths at the following time points. Consequently, at D3-4 and D5-7, Cox proportional hazards models were used. Significant univariate models were adjusted by adding invasive devices exposure durations (urinary and venous catheter, tracheal intubation) as covariates. This duration was calculated starting from patients’ admission till sampling time for each type of exposure. To allow comparison between models, hazards ratios calculated for each immune parameter were normalized to an increment from first to third quartile (interquartile range hazard ratio [IQR HR]).

For each time point, Kaplan-Meier estimations were performed. The p-value of the Log-Rank test is given. Patients were stratified in four groups based on quartiles of each immune parameter’s level calculated in all available values in all patients at all time-points.

The level of significance was set at 1%. Statistical analyses were computed with R software v3.4.4.

## Results

### Participant clinical description

Of the 1,079 patients screened during the 28-month recruitment period, 33 % (n =353) met the inclusion criteria (e-Figure 1). We included 107 septic patients, 137 trauma patients, 109 patients after elective major surgery, and 175 healthy volunteers (Table 1). A total of 65% of patients were male. The median age was 60 years [47-71]. The median SAPS II score at inclusion was 29 [20-43] and SOFA score was 5 [1-8]. At inclusion, 55% of patients presented with hemodynamic dysfunction (as illustrated by the need for vasopressor support) and 46 % with respiratory dysfunction (requiring either invasive or non-invasive mechanical ventilation). Septic and surgical patients were significantly older and had more chronic comorbidities than trauma patients. Septic patients were in a significantly more severe condition upon inclusion, with higher SAPS II and SOFA scores.

**Figure 1.**
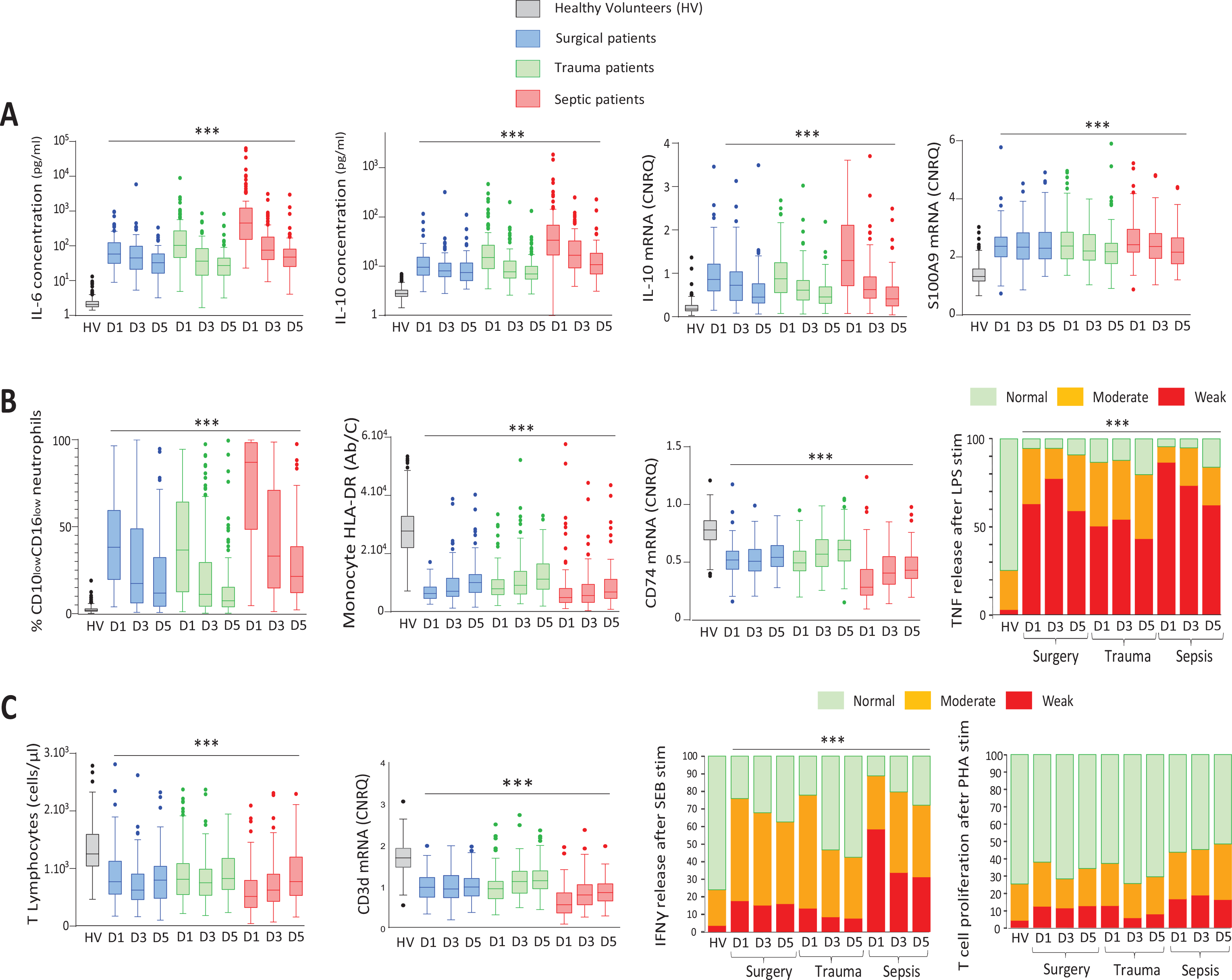
Immune monitoring in critically ill patients and healthy volunteers. Immune parameters were measured in healthy volunteers (HV, n = 175, grey boxes); surgical patients (blue boxes, n = 94 at D1-2, n = 95 at D3-4, n = 88 at D5-7), trauma patients (green boxes, n = 136 at D1-2, n = 132 at D3-4, n=119 at D5-7) and septic patients (red boxes, n = 92 at D1-2, n = 99 at D3-4, n = 94 at D5-7). **A.** Systemic pro/anti-inflammatory response **B.** Innate immune response. Ab/C = number of anti-HLA-DR antibody bound per monocyte, LPS: lipopolysaccharide. **C.** Adaptive immune response. SEB: Staphylococcus enterotoxin B, PHA: phytohemagglutinin. Results are presented as Tukey’s boxplots at each sampling time-point in each subgroup of patients or HV. For functional assays, results are presented as stacked bar plots. *Ex vivo* responses were categorized as weak (red bars, i.e response below the lowest value of the reference interval calculated in HV), moderate (orange bars, i.e. responses comprised between lowest value of reference interval and 25th percentile of HV) or normal (green bars, i.e. superior to 25th percentile of HV). Results are presented as proportions of patients in each category. Chi square or Fisher’s exact tests were used for qualitative variables’ assessment. Quantitative variables were compared with Mann-Whitney *U* test or Student t tests. *** p < 0.001 between patients and HV.

Overall, 74/353 (21%) of patients in the study developed at least one secondary infection within the first 30 days after enrolment and mortality at D30 was 5%. The mortality rate of septic patients was higher (16 %) than that of trauma and surgical patients. The incidence of secondary infections was however similar among the three studied groups of patients.

### Description of pro/anti-inflammatory, innate and adaptive immune responses over time

A detailed immune profiling was performed during the first week after admission. For each immune parameter, reference intervals were established based on results obtained in healthy volunteers (e-Table 1).

The immediate (i.e. D1-2) pro/anti-inflammatory balance after injury was characterized by the simultaneous induction of pro- (IL-6) and anti-inflammatory (IL-10) cytokines, as well as by the increased S100A9 alarmin mRNA level in the three groups of patients (Figure 1A, e-Figure 2).

**Figure 2.**
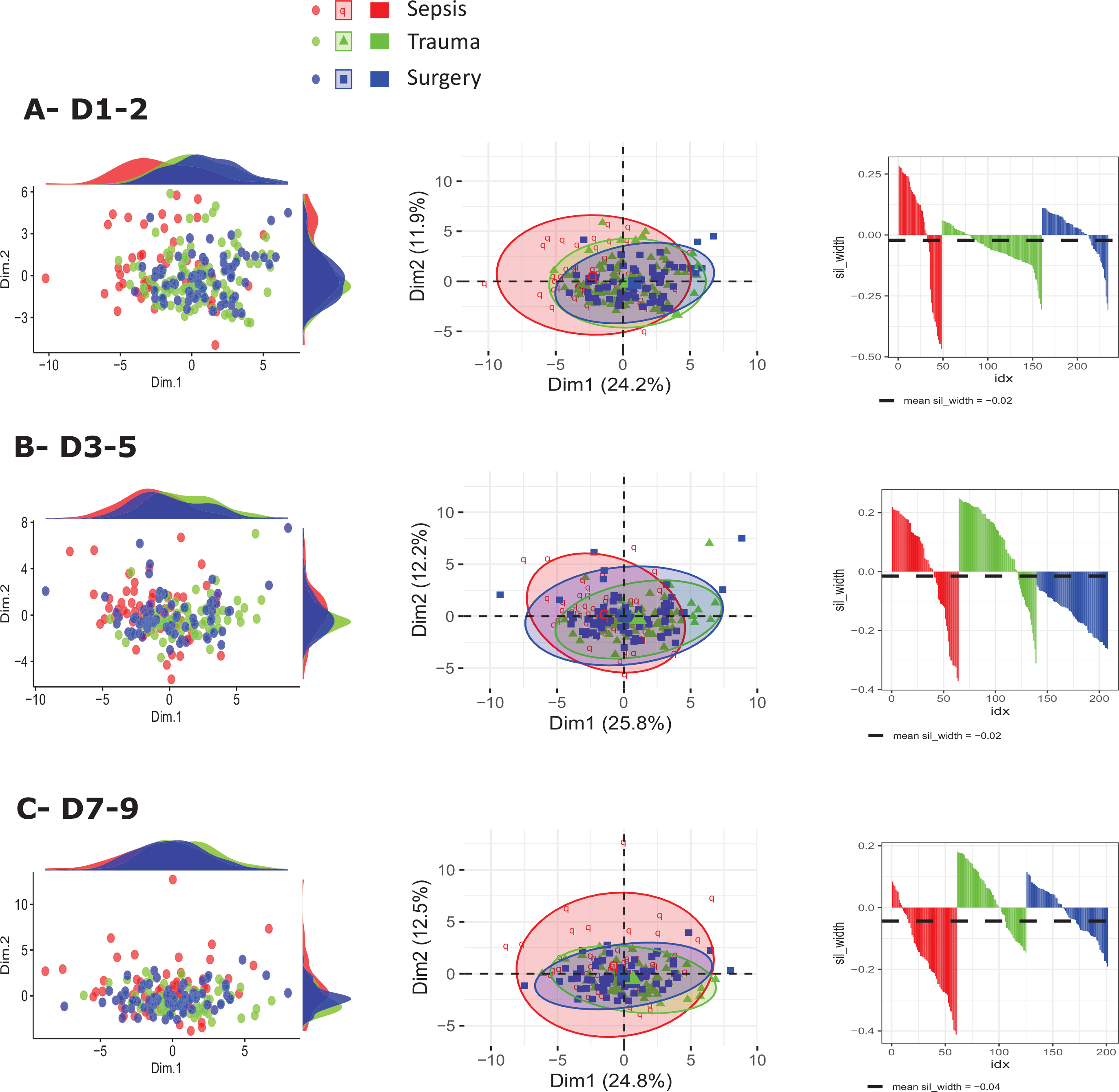
Principal component analyses of patients’ immune profiles. Principal Component Analyses (PCA) were implemented at each time-point to evaluate differences of immune profiles across subgroups. Samples from septic patients were symbolized in red (n = 48 at D1-2, n = 64 at D3-4, n = 60 at D5-7), trauma patients in green (n = 112 at D1-2, n = 75 at D3-4, n = 65 at D5-7), surgical patients in blue (n = 74 at D1-2, n = 69 at D3-4, n= 76 at D5-7). Results were presented as individual values and marginal density distributions (left graphs), individual values and confidence ellipses of PCA (middle graphs) and silhouette visualization (right graphs) for each group at each time-point (D1-2, D3-4, D5-7). For this last analysis, mean silhouettes width were mentioned.

In each group, concentrations of these cytokines and alarmins or respective mRNA were maximal at D1-2 and decreased overtime. This initial pro/anti-inflammatory response was strongly correlated with initial severity as measured by SOFA score (e-Figure 3).

**Figure 3.**
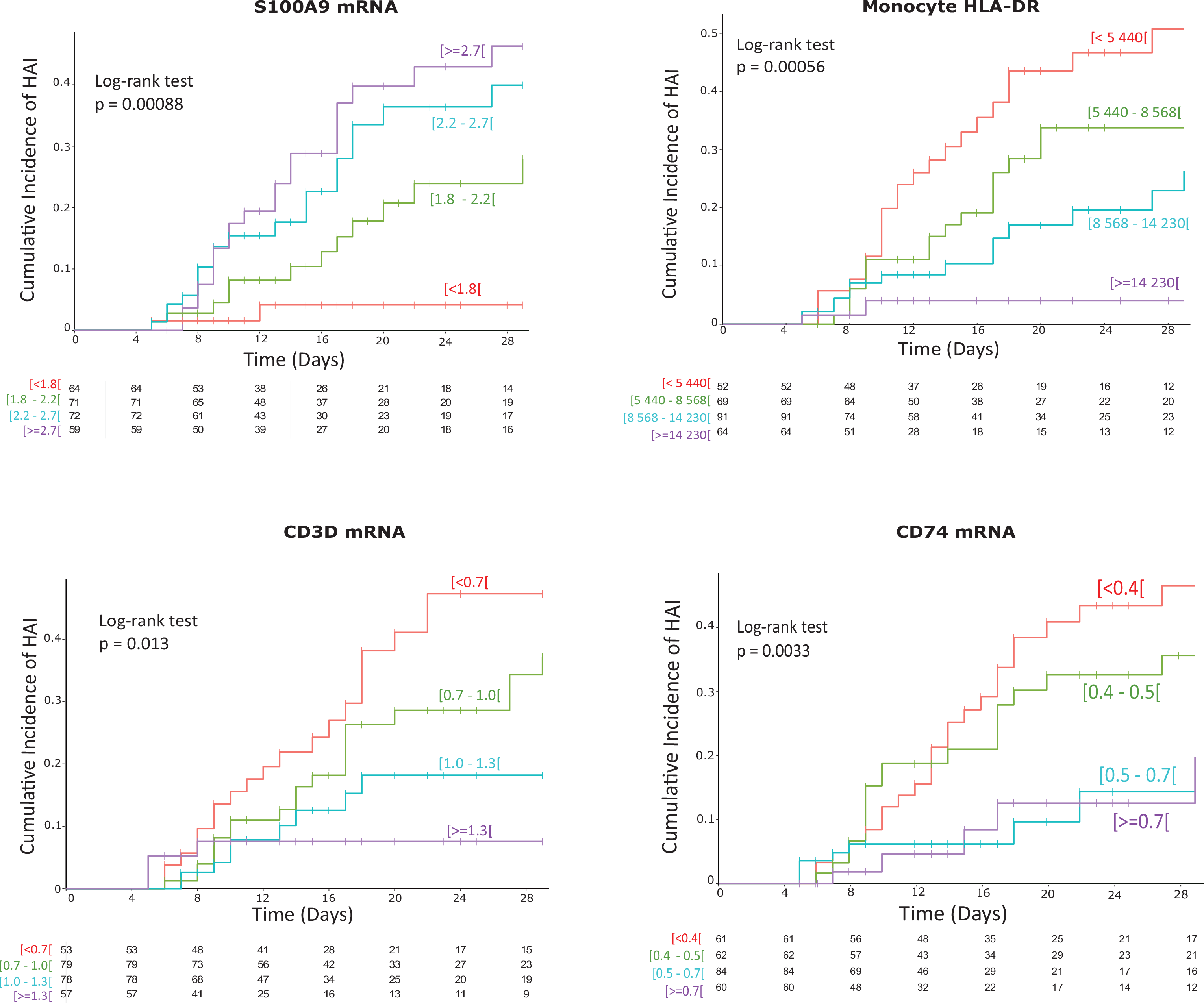
Cumulative incidence curves of secondary infections according to S100A9, HLA-DR, CD3D or CD74 levels at D5-7. Associations between S100A9 mRNA levels, HLA-DR expression on monocytes, CD3D mRNA levels and CD74 mRNA levels measured at D5-7 and occurrence of secondary infections at Day 30 (D30) were evaluated in patients (n = 225 non-infected patients and 51 patients with secondary infections). Unadjusted cumulative events for secondary infection probability at D30 were obtained with Kaplan-Meier estimations. Cumulative incidence curves are shown. Patients were stratified in four groups based on thresholds calculated as quartiles of the global distribution of mHLA-DR, S100A9, CD3D or CD74 mRNA values in the total population of patients at all available time points. The p-value of the Log-Rank test is given.

Critically ill patients concomitantly experienced altered innate immune response. This was characterized by a major rise in the number and proportion of circulating CD10^low^CD16^low^ immature neutrophils, whereas the number of circulating monocytes was not modified after injury (Figure 1B, e-Figure 2). Monocytes presented with a major decrease in MHC class II expression (decreased cell surface HLA-DR and CD74 mRNA expressions) and altered functionality (decreased *ex vivo* TNF-α production in response to LPS). These innate immune alterations were maximal at D1-2 in each group of patients and tend to normalize over the first week after injury.

Last, adaptive immune response was also strongly impacted by injury with a major decrease in the number of all lymphocyte subpopulations (Figure 1C and e-Figure 2). This was associated with decreased CD3D and CD127 mRNA levels (Figure 1C and e-Figure 2). Circulating T cells were dysfunctional as illustrated by their decreased effector responses *ex vivo* (i.e. proliferation and cytokine production after stimulation, Figure 1C and e-Figure 2).

### Impact of type of injury on immune response

To evaluate the impact of the type of injury on the immune profile of critically ill patients, an analysis of covariance (ANCOVA) was performed at each time point. To take into account the differences in age and initial severity between patients’ groups, this analysis was adjusted on age, SOFA score at D1 and Charlson score. In addition, principal component analyses (PCA) of patients’ immune profiles were established.

We observed that, except for T cell and CD4+ T cell counts measured at D5-7, none of the 30 immune markers evaluated in this analysis were significantly different between groups in ANCOVA analysis (e-Table 2). In addition, marginal density distributions and confidence ellipses of PCA presented with large overlaps between patients groups (Figure 2) and mean silhouettes width were close to zero showing the absence of any distinct subgroup clustering.

**Table 2.** Cohort description at inclusion according to development of secondary infections or mortality. Clinical description of the whole cohort and of patients who stayed alive and free of secondary infections until day 30, of patients who developed secondary health-care associated infections (HAI) and of patients who died within D30 without HAI is presented. BMI: Body Mass Index. SAPS II: Simplified Acute Physiological Score II. SOFA: Sequential Organ Failure Assessment score. D: Day. Cat: catecholamine. MV: mechanical ventilation. RRT: Renal Replacement Therapy. ICU: Intensive Care Unit. G: Giga. GCS: Glasgow Coma Scale. Hosp: hospital. ICU: Intensive Care unit. Data are presented as numbers and percentages (qualitative variables) and medians and 25th/75th percentiles (quantitative variables). Clinical and biological parameters were compared between subgroups using Kruskal-Wallis test. * until D30

| Variable | Whole cohort<br>n=353 | Alive* no HAI<br>n=263 (75%) | At least one<br>HAI<br>N=74 (21%) | Dead* no HAI<br>n=16 (5%) | P-value |
| --- | --- | --- | --- | --- | --- |
| Age (year) | 60 [47-71] | 57 [44-70] | 66 [54-72] | 75 [71-79] | < 0.001 |
| Gender (Female) | 122 (34.6 %) | 89 (33.8 %) | 28 (37.8 %) | 5 (31.3%) | 0.829 |
| BMI (kg/m2) | 25.0 [22.3-28.4] | 24.7 [22.3-27.6] | 26.3 [22.2-29.7] | 24.6 [22.8-30.6] | 0.235 |
| **Clinical scores** |  |  |  |  |  |
| Mac Cabe |  |  |  |  | 0.003 |
| Non fatal disease | 243 (68.8 %) | 192 (73.0 %) | 42 (56.8 %) | 9 (56.3 %) |  |
| Ultimately fatal disease | 98 (27.8 %) | 64 (24.3 %) | 30 (40.5 %) | 4 (25.0 %) |  |
| Rapidly fatal disease | 12 (3.4 %) | 7 (2.7 %) | 2 (2.7 %) | 3 (18.8 %) |  |
| Charlson | 1 [0-2] | 1 [0-2] | 2 [0-3] | 3 [2-3] | < 0.001 |
| SAPS II | 29 [20-43] | 26 [18-40] | 32 [22-45] | 55 [50-60] | < 0.001 |
| SOFA at D1 | 5 [1-8] | 4 [1-8] | 5 [1-8] | 11 [9-12] | < 0.001 |
| SOFA at D3-4 | 2 [0-5] | 1 [0-4] | 3 [1-6] | 10 [8-13] | < 0.001 |
| SOFA at D5-7 | 1 [0-2] | 0 [0-1] | 1 [0-4] | 8 [4-11] | < 0.001 |
| **Organ failures at D1** |  |  |  |  |  |
| Vasopressors | 193 (55 %) | 129 (49 %) | 48 (65 %) | 16 (100 %) | < 0.001 |
| D30 Cat. free days | 28 [26-29] | 29 [28-29] | 28 [25-29] | 0 [0] | < 0.001 |
| Hydrocortisone | 38 (11 %) | 14 (5.3 %) | 12 (16 %) | 12 (75 %) | < 0.001 |
| Mechanical ventilation | 164 (46 %) | 105 (40 %) | 45 (61 %) | 14 (88 %) | < 0.001 |
| D30 MV free days | 28 [22-29] | 29 [27-29] | 25 [11-29] | 0 [0] | < 0.001 |
| PaO<sub>2</sub>/FiO<sub>2</sub> | 245 [174-343] | 246 [181-358] | 265 [170-328] | 185 [135-240] | 0.044 |
| Urine output (L/day) | 1.4 [1.0-2.1] | 1.5 [1.1-2.1] | 1.4 [0.9-2.1] | 0.5 [0.1-1.3] | 0.001 |
| RRT | 61 (17 %) | 31 (12 %) | 19 (26 %) | 11 (69 %) | < 0.001 |
| D30 RRT free days | 14 [0-28] | 26 [21-28] | 13 [0-29] | 0 [0] | < 0.001 |
| Coma (GCS < 8) | 26 (7.4 %) | 19 (7.2 %) | 6 (8.1 %) | 1 (6.3 %) | 0.925 |
| **Lab values at D1-2** |  |  |  |  |  |
| pH | 7.36 [7.30-7.40] | 7.36 [7.30-7.41] | 7.36 [7.31-7.39] | 7.28 [7.24-7.38] | 0.080 |
| Lactate (mM) | 2.2 [1.7-3.3] | 2.2 [1.6-3.2] | 2.2 [1.8-3.1] | 3.2 [2.0-4.7] | 0.137 |
| Creatinine (μM) | 90 [67-142] | 87 [66-126] | 102 [70-155] | 208 [88-233] | 0.005 |
| Bilirubin (μM) | 16 [10-35] | 15 [9-35] | 21 [12-30] | 14 [12-61] | 0.628 |
| ASAT (UI/L) | 95 [49-192] | 83 [48-177] | 112 [48-179] | 208 [111-772] | 0.052 |
| Leukocytes (G/L) | 13.2 [10.2-17.3] | 12.7 [10.3-16.4] | 14.0 [10.3-18.4] | 14.6 [5.7-17.2] | 0.564 |
| Hemoglobin (g/L) | 116 [101-132] | 117 [103-132] | 114 [96-126] | 101 [87-134] | 0.137 |
| Platelets (G/L) | 205 [156-267] | 206 [162-274] | 207 [155-243] | 158 [116-223] | 0.084 |
| Neutrophils (G/L) | 10.9 [8.0-14.8] | 10.7 [7.9-14.1] | 11.8 [8.4-15.8] | 11.5 [6.7-15.9] | 0.538 |
| Lymphocytes (G/L) | 1.2 [0.8-1.8] | 1.3 [0.8-1.8] | 1.2 [0.9-1.7] | 0.6 [0.4-0.9] | 0.001 |
| Monocytes (G/L) | 0.9 [0.7-1.3] | 0.9 [0.7-1.3] | 1.1 [0.7-1.4] | 0.7 [0.6-1.1] | 0.128 |
| **Invasive device exposure** |  |  |  |  |  |
| Central line | 219 (62 %) | 145 (55 %) | 58 (78 %) | 16 (100 %) | <0.001 |
| D30 cent. line free days | 21 [3-26] | 23 [17-27] | 6 [0-18] | 0 [0] | <0.001 |
| Urinary catheter | 294 (83 %) | 213 (81 %) | 65 (88 %) | 16 (100 %) | 0.068 |
| D30 U. cath. free days | 25 [17-28] | 27 [22-28] | 16 [0-24] | 0 [0] | <0.001 |
| Tracheal intubation | 165 (47 %) | 106 (40 %) | 45 (61 %) | 14 (88 %) | <0.001 |
| D30 MV free days | 27 [19-29] | 29 [27-29] | 23 [0-29] | 0 [0] | <0.001 |
| Re-intubation rate | 12 (7.1 %) | 4 (3.6 %) | 7 (15.6 %) | 1 (7.1 %) | 0.030 |
| **Outcomes** |  |  |  |  |  |
| ICU mortality | 21 (7.0 %) | 1 (0.5 %) | 4 (6.5 %) | 16 (100 %) | <0.001 |
| D30 mortality | 20 (5.7 %) | 0 (0 %) | 4 (5.4 %) | 16 (100 %) | <0.001 |
| D90 mortality | 33 (9.4 %) | 5 (1.9 %) | 12 (16.2 %) | 16 (100 %) | <0.001 |
| D30 ICU free days | 23 [18-26] | 25 [22-27] | 18 [4-23] | 0 [0] | <0.001 |
| D30 hosp. free days | 15 [2-21] | 17 [8-22] | 2 [0-9] | 0 [0] | <0.001 |
| D30 follow-up (location) |  |  |  |  | <0.001 |
| ICU | 19 (5.7 %) | 4 (1.5 %) | 15 (21.4 %) | 0 (0 %) |  |
| Continuing care unit | 7 (2.1 %) | 2 (0.8 %) | 5 (7.1 %) | 0 (0 %) |  |
| Short-term stay | 79 (23.8 %) | 47 (17.9 %) | 32 (45.7 %) | 0 (0 %) |  |
| Long-term care | 65 (19.6 %) | 59 (22.5 %) | 6 (8.6 %) | 0 (0 %) |  |
| Home | 162 (48.8 %) | 150 (57.3 %) | 12 (17.1 %) | 0 (0 %) |  |

This analysis highlighted the absence of patients clustering according to type of injuries. Thus, based on the panel of 30 immune markers included in this study, we observed a similar regulation of the immune response in septic, surgical and trauma patients during the first week after ICU admission.

### Associations with occurrence of secondary infections

Next we evaluated the association between immune markers and secondary infections as a proxy of immune failure. Clinical and demographic characteristics at enrolment of patients with or without secondary infections and patients who died before developing such infections are presented in Table 2.

Critically ill patients who developed a secondary infection were not necessarily in a significantly more severe condition at admission than those who did not, based on SAPS II score (26 [18-40] *vs* 32 [22-45]). D90 mortality was significantly higher in the group of patients who developed at least one secondary infection (2% in the non-infected group *vs* 16% in the group with secondary infections). Patients with secondary infections were significantly more exposed to invasive medical devices, i.e. presence of a central line (78% *vs* 55%) and duration of catheterization (number of catheter-free days: 6 *vs* 23), presence of tracheal intubation (61% *vs* 40%), as well as reintubation rate (16% *vs* 4%).

In a multivariate analysis including usual clinical confounders (i.e. exposure to invasive devices), we did not observe any significant association between immune parameters measured at D1-2 and risk of secondary infection (Table 3 and e-Table 3). At D3-5, only increased S100A9 mRNA was significantly associated with increased risk of secondary infection (e-Figure 4). This suggests that, although largely deregulated compared with normal values, initial injury-induced immune response may not be a deleterious factor *per se* but rather represents an adaptive immune response to injury.

**Figure 4.**
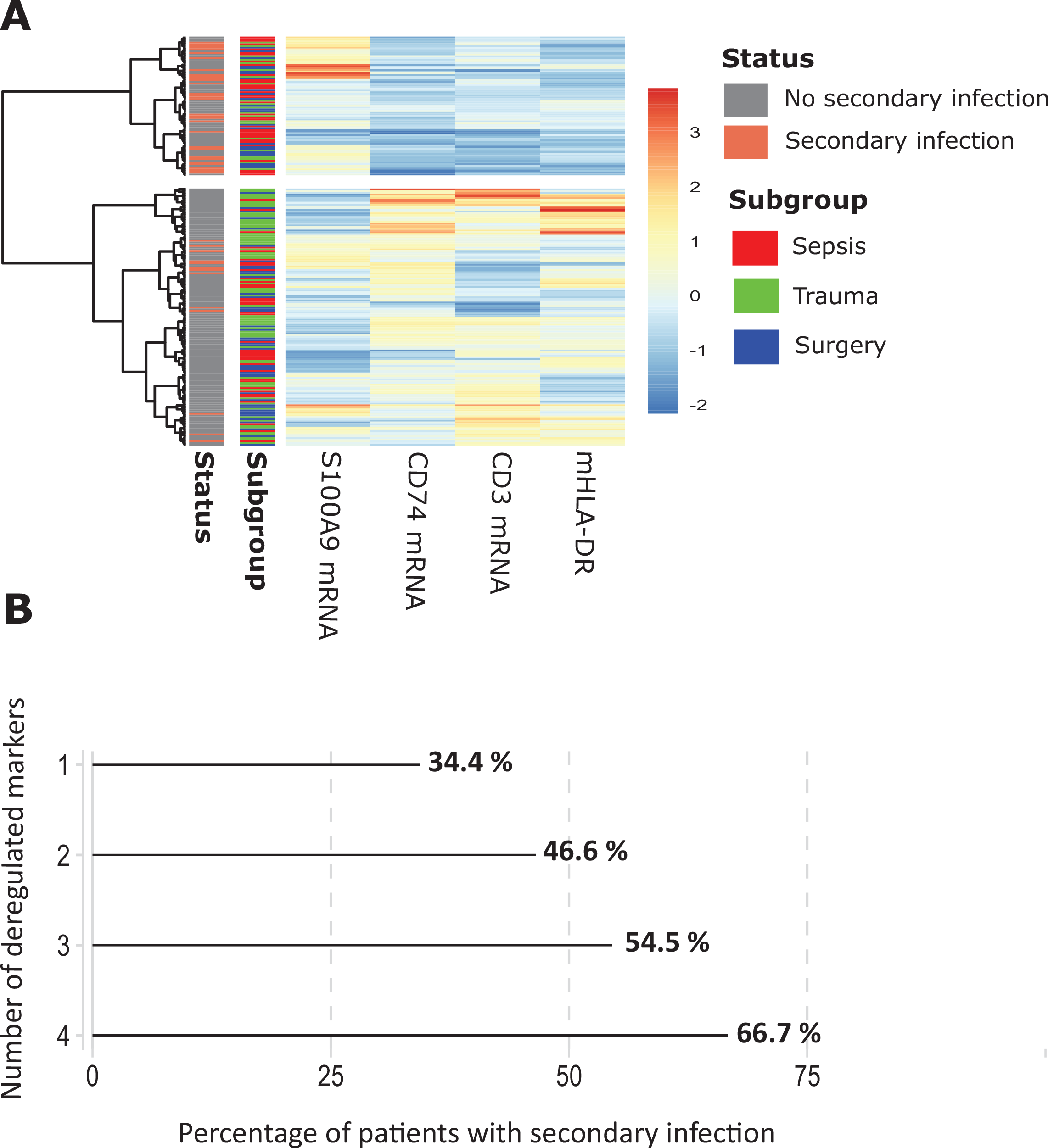
Immune profiling in critically ill patients. **A.** Hierarchical clustering of severely injured patients according to CD3D, CD74, mHLA-DR and S100A9 expressions at D5-D7 (n = 231). Patients who died without secondary infection or patients who developed a secondary infection prior to their sampling day were excluded along with outliers based on 1.5*IQR method (Tukey’s method) applied to boxcox normalized marker (according to Shapiro-Wilk normality test). Marker were centered and scaled (heatmap color scale from blue to red) and patient pairwise distance was estimated using Euclidean metric. Ward’s minimum variance method was use for patient hierarchical clustering. Resulting dendrogram (left of the heatmap) was cut into 2 clusters according to highest observed silhouette score (0.23). Patients’ status at D30 was labeled as either secondary infection (red) or no secondary infections (gray). Patients’ admission subgroups were labeled as either sepsis (red), trauma (green) or surgery (blue). **B.** Barplots of the incidence of secondary infections (x-axis) were computed according to the number of dysregulated immune markers among CD3D, CD74, mHLA-DR and S100A9 expressions at D5-D7 (ranging from 1 to 4, y-axis, n = 257 patients among whom 132 had no dysregulated marker). The incidences were indicated as percentages on the right of each bars. Dysregulation threshold was set as either Q1 or Q3 according to the furthest one from median value of healthy donors. Patients who died without secondary infection or patients who developed a secondary infection prior to their sampling day were excluded.

**Table 3.**
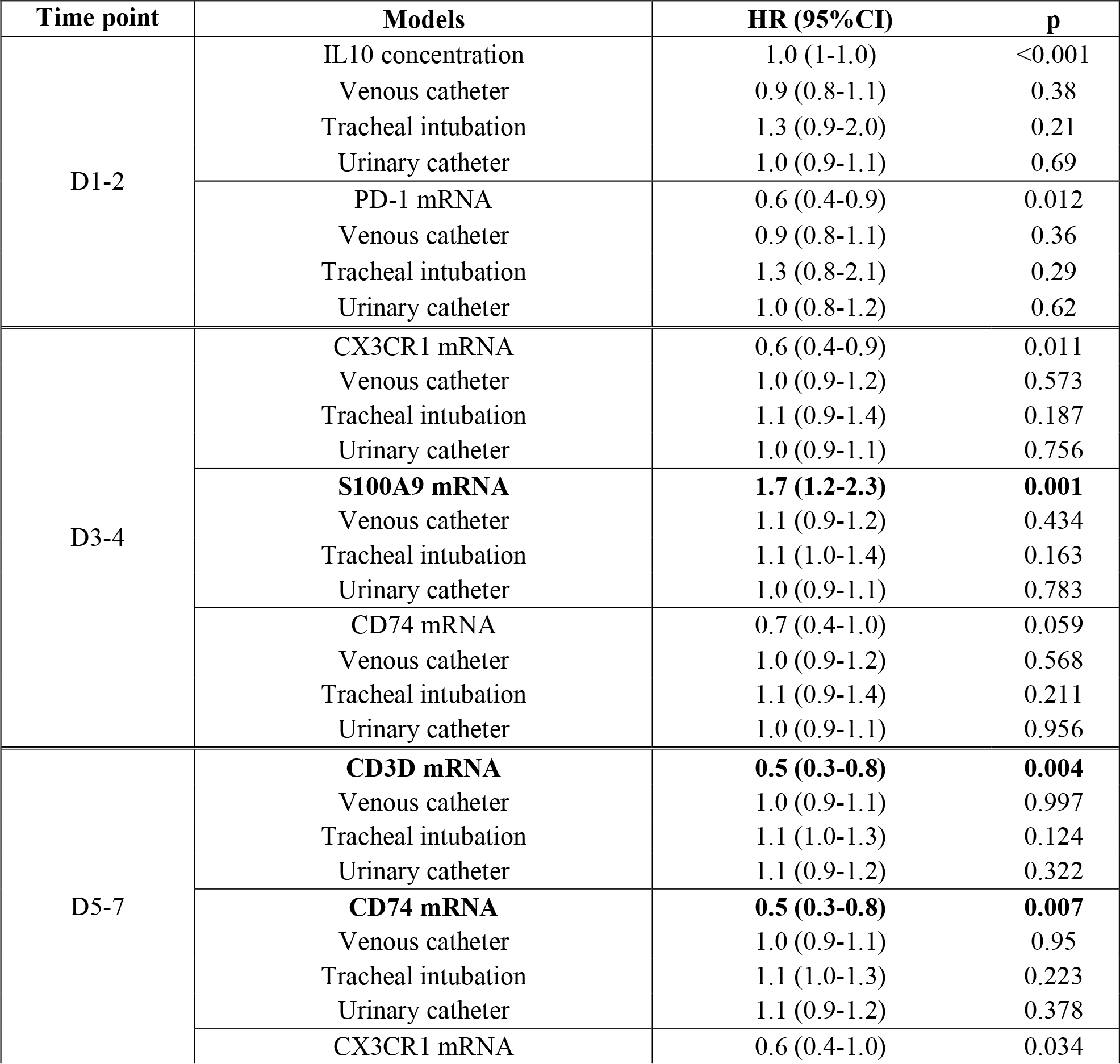
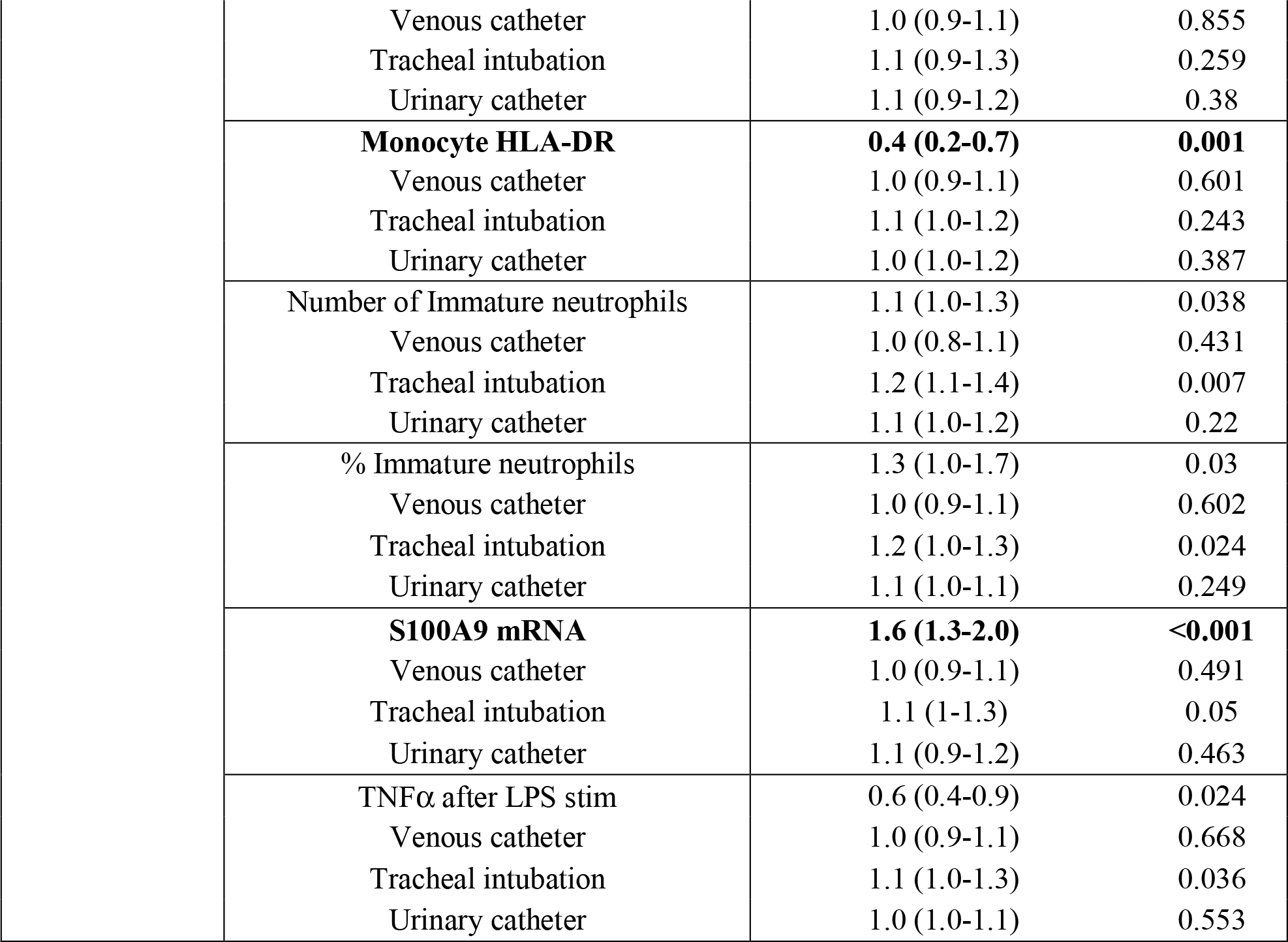
Multivariate analyses of associations between immune parameters and secondary infections at D30. Three hundred and fifty three critically ill patients were prospectively included among whom 74 patients developed at least one secondary infection within 30 days (D) and 16 died. The associations between D30 secondary infection and immune parameters (with death as competitive risk) were performed by implementing univariate Fine and Gray models at D1-2. Due to the insufficient number of deaths at the following time points, Cox proportional hazards models were used at D3-4 and D5-7. Significant univariate models were adjusted by adding invasive devices exposure durations (urinary and venous catheter, tracheal intubation) as covariates. This duration was calculated starting from patients’ admission till sampling time for each type of exposure. To allow comparison between models, hazards ratios (HR) calculated for each immune parameter were normalized to an increment from first to third quartile. Significant associations between immune parameters and secondary infections were considered if p values ≤0.05 and HR 95% confidence intervals did not include 1. These are highlighted in bold.

| Time point | Models | HR (95%CI) | p |
| --- | --- | --- | --- |
| D1-2 | IL10 concentration | 1.0 (1-1.0) | <0.001 |
|  | Venous catheter | 0.9 (0.8-1.1) | 0.38 |
|  | Tracheal intubation | 1.3 (0.9-2.0) | 0.21 |
|  | Urinary catheter | 1.0 (0.9-1.1) | 0.69 |
|  | PD-1 mRNA | 0.6 (0.4-0.9) | 0.012 |
|  | Venous catheter | 0.9 (0.8-1.1) | 0.36 |
|  | Tracheal intubation | 1.3 (0.8-2.1) | 0.29 |
|  | Urinary catheter | 1.0 (0.8-1.2) | 0.62 |
| D3-4 | CX3CR1 mRNA | 0.6 (0.4-0.9) | 0.011 |
|  | Venous catheter | 1.0 (0.9-1.2) | 0.573 |
|  | Tracheal intubation | 1.1 (0.9-1.4) | 0.187 |
|  | Urinary catheter | 1.0 (0.9-1.1) | 0.756 |
|  | **S100A9 mRNA** | **1.7 (1.2-2.3)** | **0.001** |
|  | Venous catheter | 1.1 (0.9-1.2) | 0.434 |
|  | Tracheal intubation | 1.1 (1.0-1.4) | 0.163 |
|  | Urinary catheter | 1.0 (0.9-1.1) | 0.783 |
|  | CD74 mRNA | 0.7 (0.4-1.0) | 0.059 |
|  | Venous catheter | 1.0 (0.9-1.2) | 0.568 |
|  | Tracheal intubation | 1.1 (0.9-1.4) | 0.211 |
|  | Urinary catheter | 1.0 (0.9-1.1) | 0.956 |
| D5-7 | **CD3D mRNA** | **0.5 (0.3-0.8)** | **0.004** |
|  | Venous catheter | 1.0 (0.9-1.1) | 0.997 |
|  | Tracheal intubation | 1.1 (1.0-1.3) | 0.124 |
|  | Urinary catheter | 1.1 (0.9-1.2) | 0.322 |
|  | **CD74 mRNA** | **0.5 (0.3-0.8)** | **0.007** |
|  | Venous catheter | 1.0 (0.9-1.1) | 0.95 |
|  | Tracheal intubation | 1.1 (1.0-1.3) | 0.223 |
|  | Urinary catheter | 1.1 (0.9-1.2) | 0.378 |
|  | CX3CR1 mRNA | 0.6 (0.4-1.0) | 0.034 |

|  |  |  |
| --- | --- | --- |
| Venous catheter | 1.0 (0.9-1.1) | 0.855 |
| Tracheal intubation | 1.1 (0.9-1.3) | 0.259 |
| Urinary catheter | 1.1 (0.9-1.2) | 0.38 |
| **Monocyte HLA-DR** | **0.4 (0.2-0.7)** | **0.001** |
| Venous catheter | 1.0 (0.9-1.1) | 0.601 |
| Tracheal intubation | 1.1 (1.0-1.2) | 0.243 |
| Urinary catheter | 1.0 (1.0-1.2) | 0.387 |
| Number of Immature neutrophils | 1.1 (1.0-1.3) | 0.038 |
| Venous catheter | 1.0 (0.8-1.1) | 0.431 |
| Tracheal intubation | 1.2 (1.1-1.4) | 0.007 |
| Urinary catheter | 1.1 (1.0-1.2) | 0.22 |
| % Immature neutrophils | 1.3 (1.0-1.7) | 0.03 |
| Venous catheter | 1.0 (0.9-1.1) | 0.602 |
| Tracheal intubation | 1.2 (1.0-1.3) | 0.024 |
| Urinary catheter | 1.1 (1.0-1.1) | 0.249 |
| **S100A9 mRNA** | **1.6 (1.3-2.0)** | **<0.001** |
| Venous catheter | 1.0 (0.9-1.1) | 0.491 |
| Tracheal intubation | 1.1 (1-1.3) | 0.05 |
| Urinary catheter | 1.1 (0.9-1.2) | 0.463 |
| TNFα after LPS stim | 0.6 (0.4-0.9) | 0.024 |
| Venous catheter | 1.0 (0.9-1.1) | 0.668 |
| Tracheal intubation | 1.1 (1.0-1.3) | 0.036 |
| Urinary catheter | 1.0 (1.0-1.1) | 0.553 |

However, at D5-7, persistent increase in S100A9 mRNA level, decrease in CD3D mRNA level and altered monocyte response (CD74 and HLA-DR expression) were associated with increased risk of secondary infections regardless of the duration of exposure to invasive devices (Figure 3). Thus, the presence of profound immune alterations in some patients at the end of the first week after injury is an independently risk factor for an increased risk of secondary infections. Markers representing either pro/anti-inflammatory, innate and adaptive immune responses were significantly associated with increased risk of infection. In this subgroup of patients, persistence of such immune alterations represents a maladaptive response to injury associated with deleterious outcomes.

### Immune profiling

Finally, we evaluated the importance to combine these four markers reflecting complementary aspects of the immune response to better appreciate patients’ immune status.

Unsupervised clustering based on S100A9, CD3D, CD74 mRNA levels and mHLA-DR expression at D5-7 led to the separation of the cohort in two clusters (Figure 4A). Patients included in cluster 1 presented with high S100A9 mRNA and low CD3D, CD74 mRNA and mHLA-DR whereas patients from cluster 2 expressed low S100A9 but high CD3D, CD74 and mHLA-DR.

Septic, trauma and surgical patients were included in each cluster therefore confirming the similarity of the immune response described above. Cluster 1 was very significantly enriched with patients with secondary infections compared with cluster 2 (40% versus 9%, p<0.001, chi-squared test).

Finally, based on threshold used to draw Kaplan-Meier curves, we calculated the percentages of patients with secondary infection depending on the number of deregulated markers. We observed a parallel increase between the proportion of patients with secondary infection and the number of deregulated immune marker (Figure 4B). For example, 34.4% of patients with one deregulated immune function developed a secondary infection whereas almost 70% of patients with 4 deregulated markers presented a secondary infection. This indicates that the monitoring of a combination of markers reflecting complementary aspects of the immune response may be more appropriate to monitor severely injured patients immune status and capture their heterogeneity.

## Discussion

The description of the immune response after injury has been the focus of an increasing number of observational clinical studies within the last ten years 20. While this research effort has been key in demonstrating the central role of the immune response in the pathophysiology of severe injuries such as sepsis, the description of the immune profile induced by injury remains incomplete20. Indeed, most studies were limited to the evaluation of a restricted number of immune parameters, using a limited number of techniques in a single group of patients. In addition, except for some large scale transcriptomic studies^21^, most studies included small numbers of patients due to the difficulty of ensuring standardization over time of complex immune measurements such as flow cytometry or functional testing. Finally, in most studies, normal values for the measured immune parameters were missing due to the lack of inclusion of age and sex-matched groups of healthy volunteers processed concomitantly with patients. Thus, as highlighted in a recently published opinion paper by the European Group on Immunology of Sepsis, we still lack a clear blueprint of the immune response to injury^20^.

In the REALISM study, we defined a panel of 30 immune markers assessing pro/anti-inflammatory, innate and adaptive immune responses. Techniques were selected to reflect complementary aspects of the immune responses (i.e. cell numbers, phenotype, transcriptome and function as well as circulating concentrations of cytokines), and to be easy-to-measure and available in most routine labs (i.e. cellular phenotyping, functional testing, protein and mRNA measurements). Measurements procedures were standardized to ensure results reproducibility within this study but also eventually between studies. In addition, we calculated for each immune parameter the reference interval for normal values. For example, we defined that the normal values for mHLA-DR expression in the general adult population were comprised between 13 500 and 45000 Ab/C and were independent of age and sex. Markers of inflammation (plasma IL-6 and IL-10 levels, proportion of immature neutrophils) were almost undetectable in healthy volunteers’ blood. We observed that the reference intervals of some immune parameters such as the measurement of T lymphocyte proliferation were large. In addition to the wide inter-individual heterogeneity, this probably reflects the technical variability of such non-automated and mostly manual functional tests. Thus the panel of markers chosen in this study was robust and reliable to evaluate complementary aspects of pro/anti-inflammatory, innate and adaptive immune responses in a large cohort of severely injured patients.

Using this immunomonitoring panel, our results showed that the immune response induced by injury is a rather universal phenomenon which does not strongly depend on the type of injury (infectious *vs* sterile). This is in agreement with previous studies, which described in different cohorts the regulation of specific markers such as decreased mHLA-DR expression or absolute lymphocyte counts after ICU admission^11-13^. However, the comparison between different earlier studies is difficult, as they comprise patients recruited and measured differently. In contract, the results of our study show for the first time in a prospective study including a large number of septic, trauma and surgical patients that, when concomitantly measured with standardized tests, aspects of pro/anti-inflammatory, innate and adaptive immune responses were similarly regulated after injury. Indeed, binding of pathogen-associated molecular patterns (PAMPs) and danger-associated molecular patterns (DAMPs) to their common receptors induces shared intracellular signaling pathways leading to the activation of similar effector immune response and compensatory immune mechanisms 22. This allows to define the features of a common injury induced immune signature.

Starting from D1-2 and over the course of the disease, pro- and anti-inflammatory cytokine release and alterations of innate and adaptive immune responses were observed. This is in accordance with the study by Timmermans *et al*. who showed, in a cohort of trauma patients, that an anti-inflammatory phenotype could be detected at the trauma scene, i.e. within 30 min after injury 12. Thus, very early after injury, induction of the pro-inflammatory effector response was associated with the concomitant development of regulatory mechanisms to protect the host from such an overwhelming immune response.

Although the injury-induced immune response was at its highest intensity at D1-2 and while pro- and anti-inflammatory cytokine responses were strongly correlated with initial severity, no association between the immune parameters measured at this time point and increased risk of secondary infections was observed. This illustrates that, although extensive, the initial injury-induced immune response should not necessarily be seen as a deleterious factor *per se* but rather represents an adaptive response to the injury aimed at controlling the insult and protecting the host from this massive immune response. Hence, this represents the physiologic adaptive injury-induced immune response to readjust immune homeostasis.

After this initial adaptive injury-induced immune response, the immune profile of critically ill patients tends to improve over time to return homeostasis. However, at the end of the first week after injury, the presence of immune dysfunctions in a subgroup of patients was associated with an increased risk of secondary infections independently of usual confounding factors (exposure to invasive medical devices). At this time point, the persistence of injury-induced immune alterations thus represents an independent risk factor of deleterious outcome in critically ill patients. Thus, this persistent dysregulated immune profile could not be considered as part of the physiological response to injury but rather as a maladaptive evolution of the immune response in a subgroup of patients which causative mechanisms remain to be identified.

This maladaptation of the immune response aligns with the previous descriptions of the increased risk of viral reactivations^23^, fungal infections^24^ or secondary infections with weakly virulent pathogens^25^ in critically ill patients which may all be considered as reflecting acquired immune dysfunction. Overall, the combination of such immune alterations and of increased risk of secondary infections is consistent with the general definition of immunosuppression^26^. In the particular context of critically ill patients, the term delayed injury-acquired immunodeficiency may be more appropriate.

Interestingly, it is the non-resolution of complementary aspects of injury-induced immune alterations but not hyper-activation of the immune response which associated with increased risk of secondary infections. More specifically, persistently altered T lymphocyte (decreased CD3D mRNA) and monocyte responses (decreased HLA-DR expression, CD74 mRNA level and TNF production *ex vivo* in response to LPS), but not increased IL-6 levels, were markedly associated with increased risk of secondary infections although this pro-inflammatory cytokine response was strongly dysregulated in patients. One exception is the observation of the significant association between the elevated S100A9 alarmin concentration at D5-7 and increased risk of secondary infection as previously described^27^. However, as S100A9 has been proposed to participate in myeloid-derived suppressor cell immunosuppressive functions, this association could reflect the delayed increase of these immunoregulatory cells in patients^28^. This is in accordance with the concept of Persistent Inflammation, Immunosuppression, and Catabolism Syndrome (PICS) recently described in sepsis^29^. The exploration of the link between persistent inflammation, injury-acquired immunodeficiency and long-term deleterious outcome in critically ill patients is a research subject of major interest which deserves dedicated investigations.

This also highlights that, as opposed to the follow-up of a single parameter, the monitoring of severely injured patients should be performed through the combined use of a common set of markers. In addition, the concomitant monitoring of immune markers reflecting complementary aspects of the immune response improved the capacity to identify subgroups of patients at high risk of secondary infections. Thus a combination of markers reflecting pro/anti-inflammatory, innate and adaptive immune responses should be followed to capture the overall facets of immune alteration and the heterogeneity of cohorts of severely injured patients. Based on this paradigm, specific markers and combination algorithm should now be developed.

Our results support the rationale for immune intervention and implementation of specific care bundles in an identifiable subset of critically ill patients to restore their immune response and reduce risk of secondary infections. Based on our data, we suggest that such treatment could be given after initial insult in patients diagnosed with an injury-acquired immunodeficiency. In daily clinical practice we lack early predictive clinical sign of immune alteration in order to identify patients with the most severe and persistent immune alterations requiring treatment. In the future, however, our data reinforce that detailed monitoring of the host immune response will allow the development of personalized therapies that target injury-acquired immunodeficiency to reduce secondary infections and improve survival.

Our study has limitations. First, the use of a single center which was chosen to ensure standardization and reproducibility of the large number of measured immune parameters could limit the extrapolation of our results to other sites. However, our results are consistent with

previously described observations and were obtained with standardized techniques (e.g. mHLA-DR, circulating lymphocyte count). Second, although extensive, the immune monitoring panel chosen in this study was not exhaustive. In particular, no multi-omics approaches were performed. Again, this was to guarantee the standardization and reproducibility of the results that were generated on a day-by-day basis in each patient with routinely used immune tests. Finally, the comparative performances of the individual immune parameter in predicting deleterious outcomes were not evaluated and the best combination of markers to predict death or secondary infections or to diagnose injury-induced immunodeficiency syndrome was not identified. This will be the objective of a dedicated study.

In conclusion, through an extensive immune monitoring approach incorporating data from flow cytometry, functional assays, protein- and mRNA-level measurements, we describe for the first time the injury-induced immune signature. Our results show similar dynamics of the host immune response after three different types of injury, which begin with a universal initial adaptive stress immune response followed by a maladaptive injury-acquired immunodeficiency syndrome. Furthermore, this understanding provides insight into how, based on detailed immune monitoring, delayed administration of immune adjuvant therapy and the use of specific care bundles could be used to improve outcomes in the critically ill patients.

## Data Availability

Data from REALISM study are available upon reasonable request.

## Acknowledgments

The authors would like to thank: i) the clinical research assistants for their great help in the screening and sampling of patients as well as in the preparation of case report forms, ii) the technical staff from the Edouard Herriot Hospital-bioMérieux joint research unit and BIOASTER for their dedicated involvement in samples processing and iii) the team of biostatisticians led by S Blein for their great help in data analysis iv) Sophie Abblot from bioMérieux for her great help in paper editing. The authors would also like to acknowledge the important contribution and motivation of the medical and paramedical staff of the Anesthesia and Critical Care Medicine Department of the Edouard Herriot Hospital. Finally, the authors would like to thank the patients and their families for supporting this study.

## Abbreviation list

Ab/C: number of antibody bound per cell
ANCOVA: analysis of covariance
D: day
DAMP: danger associated molecular pattern
HLA-DR: human leukocyte antigen-DR
HR: hazard ratio
IL: interleukine
IQR: interquartile range
ICU: intensive care unit
ISS: injury-severity score
LPS: lipopolysaccharide
LOS: length of stay
PCA: principal component analysis
PAMP: pathogen-associated molecular pattern
SAPS II: simplified acute physiological score II
SOFA: sequential organ failure assessment score

## Authors contributions

FV, JT, SB, PC, LKT, CT, LQ, FR, PL, CV, YB, BD, OM, TG, CT, VM, AP, GM, TR were involved in the study design and data analysis. FV, JT, SB, BM, MLR, BC, PC, FR, CV, VM, AP, GM, TR contributed to the acquisition of data and data analysis. JT and FV drafted the manuscript and all authors were involved in the revision of the manuscript and approved the final version.

## Supplementary Material

### Legends to Supplementary Figures

**e-Figure 1.**
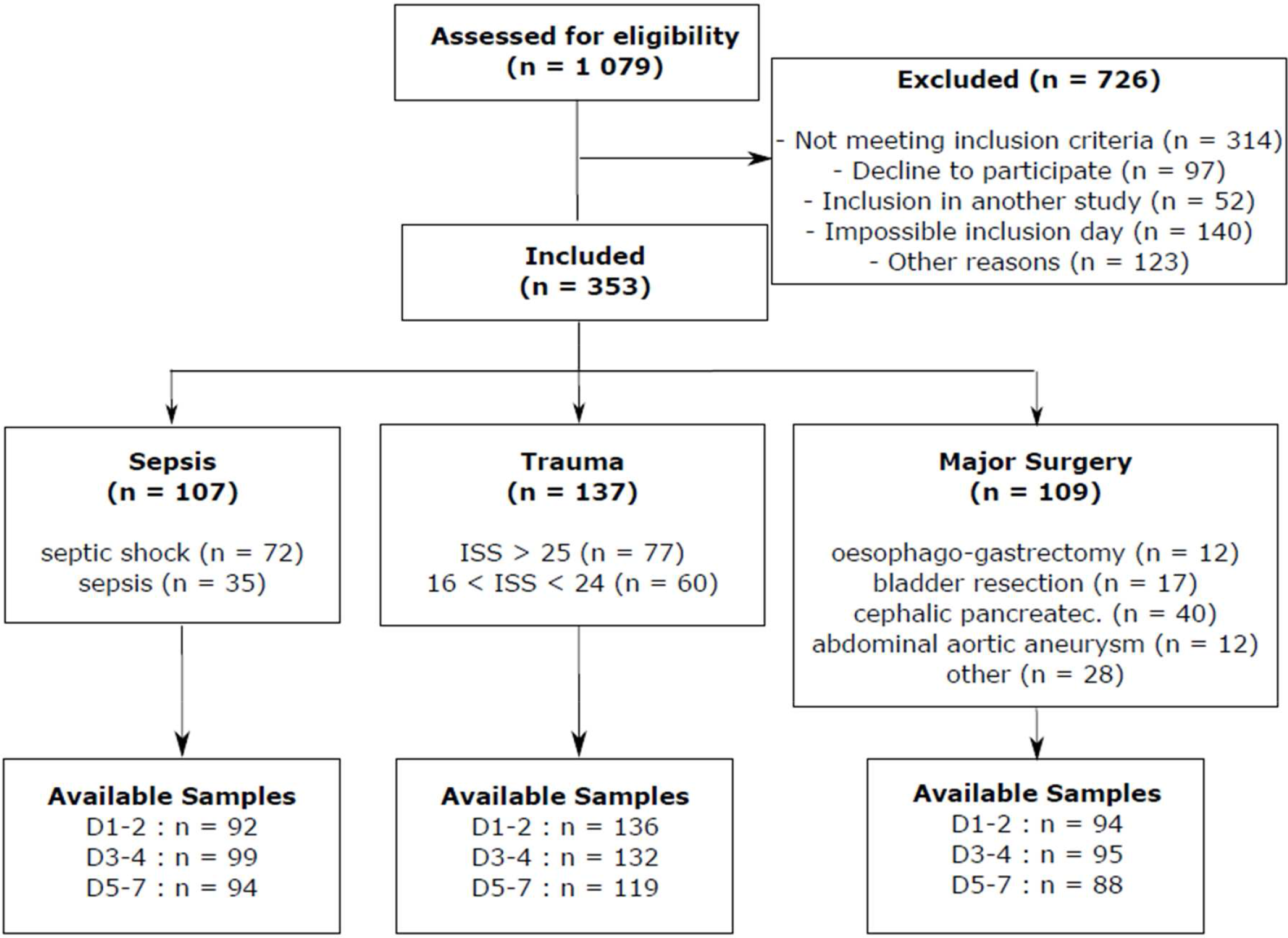
Study flow diagram. ISS: Injury Severity Score. Cephalic pancreatec: cephalic pancreaticoduodenectomy.

**e-Figure 2.**
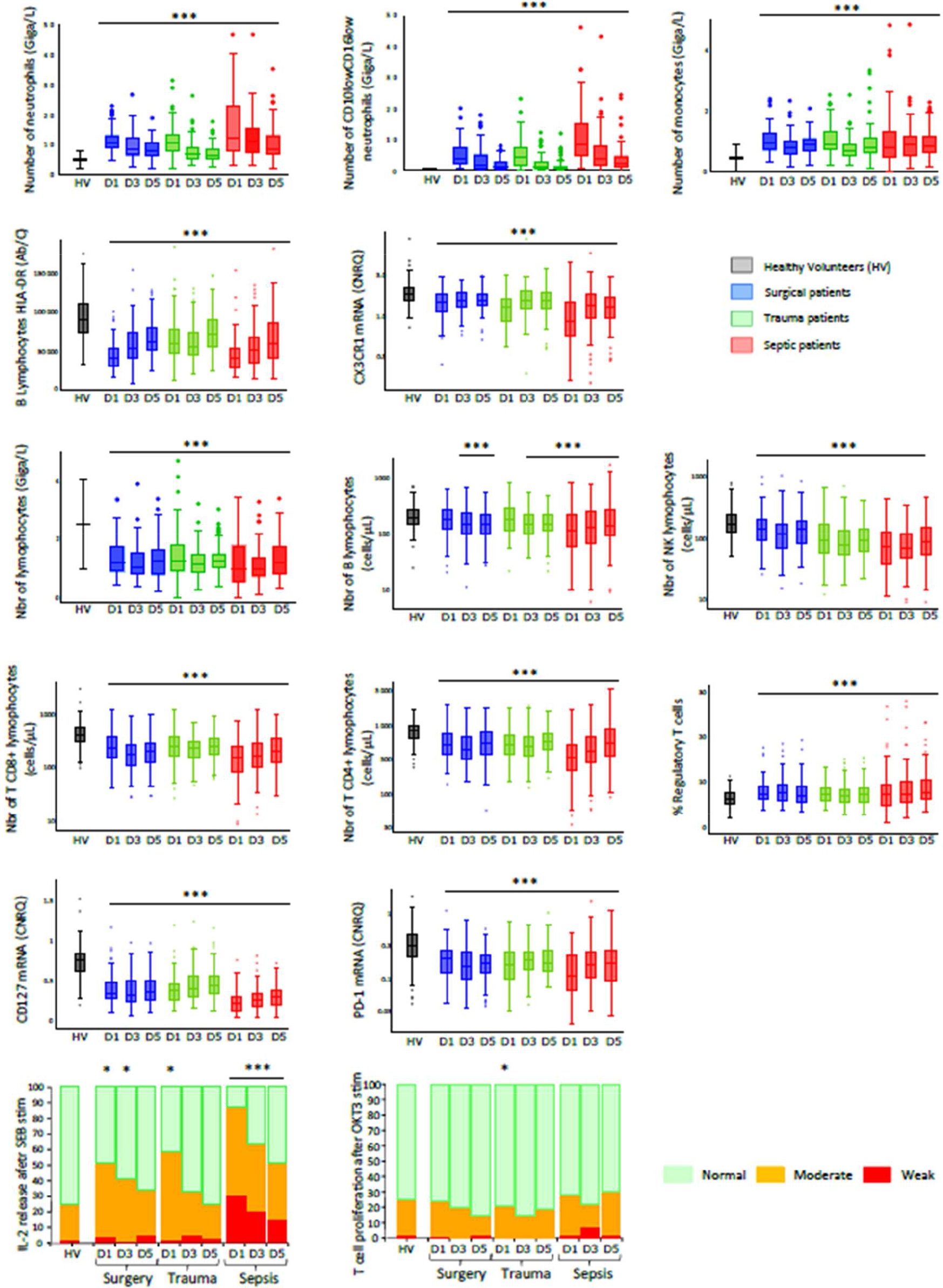
Immune monitoring in critically ill patients and healthy volunteers. Immune parameters were measured in healthy volunteers (HV, n = 175, grey boxes); surgical patients (blue boxes) at day 1-2 (D1, n = 94), D3-4 (D3, n = 95) and D5-7 (D5, n = 88); trauma patients (green boxes) at D1-2 (n = 136), D3-4 (n = 132) and D5-7 (n=119) and septic patients (red boxes) at D1-2 (n = 92), D3-4 (n = 99) and D5-7 (n = 94). Ab/C: number of anti-HLA-DR antibody bound per B lymphocyte. CNRQ: Calibrated Normalized Relative Quantity. Results are presented as Tukey’s boxplots at each sampling time-point in each subgroup of patients or HV. Samples with values outside the 1.5 interquartile ranges are presented as individual dots. Interleukin-2 (IL-2) release after Staphylococcus enterotoxin B (SEB) stimulation of whole blood and T cell proliferation after anti-CD3 antibody (OKT3) stimulation of mononuclear cells were measured. Results are presented as stacked bar plots. *Ex vivo* responses were categorized as weak (red bars, i.e response below the lowest value of the reference interval calculated in HV), moderate (orange bars, i.e. responses comprised between lowest value of reference interval and 25^th^ percentile of HV) or normal (green bars, i.e. superior to 25^th^ percentile of HV). Results are presented as proportions of patients in each category. Chi square or Fisher’s exact tests were used for qualitative variables’ assessment. Quantitative variables were compared with Mann-Whitney *U* test or Student t tests. * p < 0.05 and *** p < 0.001 between patients and HV.

**e-Figure 3.**
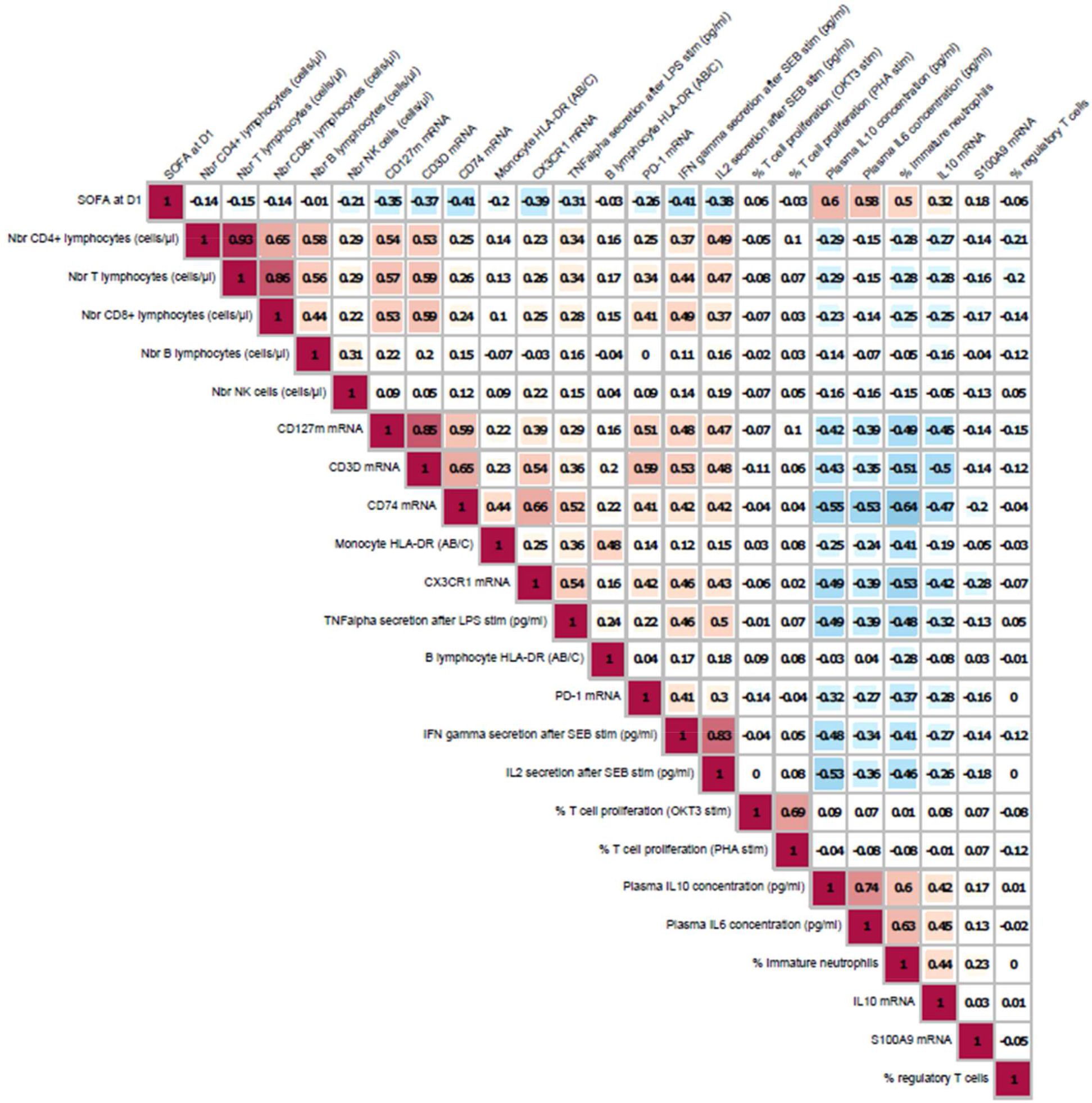
Correlations between immune parameters and SOFA at Day 1. Correlations between SOFA score at Day 1 (D1) and immune parameters measured at D1-2 in critically ill patients were evaluated. For all pairs of immune parameters, Spearman’s rho correlation coefficients were computed and summarized in a correlation matrix. The strengths of the correlations were color-coded by different shades of red for positive correlations and blue for negative correlations. Unsupervised clustering of the immune parameters was performed based on similarities between correlation coefficients. Correlation coefficients were considered as clinically relevant if > 0.5.

**e-Figure 4.**
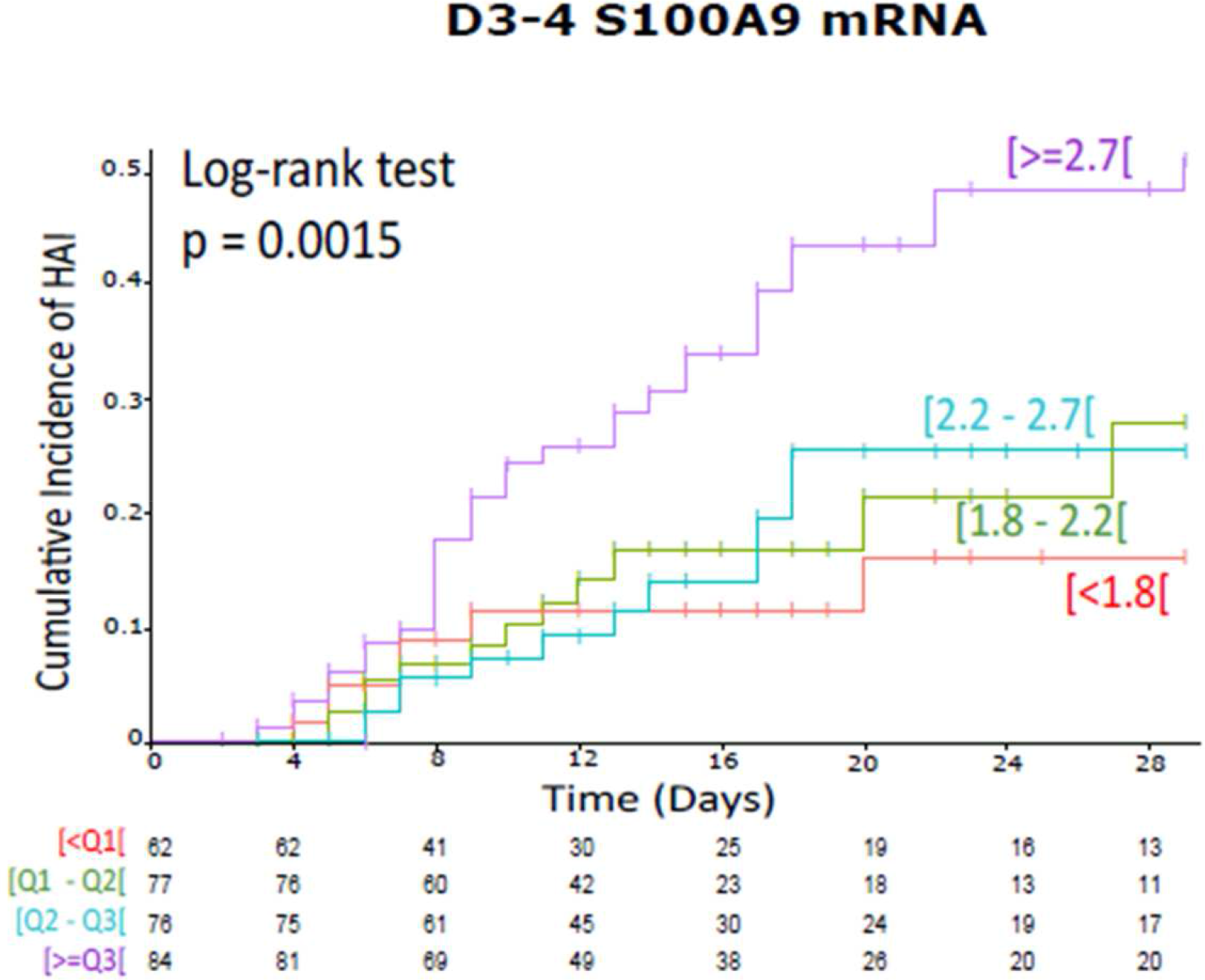
Cumulative incidence curves of secondary infections according to S100A9 mRNA expression at D3-4. The associations between S100A9 mRNA levels measured at D3-4 and occurrence of secondary infections at Day 30 (D30) were evaluated in patients (n = 225 non-infected patients and 51 patients with secondary infections). Unadjusted cumulative events for secondary infection probability at D30 were obtained with Kaplan-Meier estimations. Patients were stratified in 4 groups based on thresholds calculated as quartiles of the global distribution of S100A9 mRNA levels in the total population of patients at all available time points. The p-value of the Log-Rank test is given.

**e-Table 1.** Immune profiling panel. Magnitudes of pro and anti-inflammatory cytokine productions; phenotypic, transcriptional and functional profiles of innate (neutrophils, monocytes) and adaptive (lymphocytes) cellular responses were concomitantly assessed in a cohort of 353 critically ill patients including septic, trauma and surgical patients and 175 healthy volunteers. For each immune parameter, reference intervals were calculated based on 2.5th and 97.5th percentiles of healthy volunteers’ values. FCM = Flow CytoMetry. CNRQ = Calibrated Normalized Relative Quantity. Ab/C = number of Antibodies bound per Cells. LPS = LipoPolySaccharide. SEB = Staphylococcal Enterotoxin B. OKT3 = monoclonal antibody directed against CD3. PHA = PhytoHaemAgglutinin

| Immune Function | Marker | Type of marker | Expression unit | Assay Method | Reference interval |
| --- | --- | --- | --- | --- | --- |
| **Pro / Anti-inflammatory responses** | IL-6 concentration | Soluble marker | pg/mL | ELISA | [1 - 4] |
|  | IL-10 concentration | Soluble marker | pg/mL | ELISA | [2 - 4] |
|  | IL-10 mRNA | Transcriptomic | CNRQ | qRT-PCR | [0.04 - 0.40] |
|  | S100A9 mRNA | Transcriptomic | CNRQ | qRT-PCR | [0.80 - 2.09] |
| **Innate immune response** | Neutrophil count | Cellular marker | Giga/L | Hemocytometer | [1.8 – 7.5] |
|  | CD16<sup>low</sup>CD10<sup>low</sup> neutrophils | Cellular marker | Giga/L | FCM | [0 – 0.4] |
|  | CD16<sup>low</sup>CD10<sup>low</sup> neutrophils | Cellular marker | % among neutrophils | FCM | [0- 5] |
|  | Monocyte count | Cellular marker | Giga/L | Hemocytometer | [0.2 – 0.9] |
|  | HLA-DR on monocytes | Cellular marker | Ab/C | FCM | [13 500 – 45 000] |
|  | HLA-DR on B lymphocytes | Cellular marker | Ab/C | FCM | [47 500 – 151 000] |
|  | CD74 mRNA | Transcriptomic | CNRQ | qRT-PCR | [0.55 - 1.03] |
|  | CX3CR1 mRNA | Transcriptomic | CNRQ | qRT-PCR | [0.97 - 2.91] |
|  | TNFα release after LPS stimulation | Functional test | pg/mL | ELISA | [1 874 – 11 524] |
| **Adaptive Immune response** | Total lymphocyte count | Cellular marker | Giga/L | Hemocytometer | [1 – 4] |
|  | T Lymphocyte count | Cellular marker | cells/μL | FCM | [594 – 2 156] |
|  | T CD4<sup>+</sup> lymphocyte count | Cellular marker | cells/μL | FCM | [365 – 1 345] |
|  | T CD8<sup>+</sup> lymphocyte count | Cellular marker | cells/μL | FCM | [158 - 857] |
|  | B lymphocyte count | Cellular marker | cells/μL | FCM | [82 - 374] |
|  | NK cell count | Cellular marker | cells/μL | FCM | [634- 361] |
|  | CD4<sup>+</sup>CD25<sup>high</sup>CD127<sup>Low</sup> regulatory T cells | Cellular marker | % among CD4+ lymphocytes | FCM | [3- 10] |
|  | CD3D mRNA | Transcriptomic | CNRQ | qRT-PCR | [1.08 - 2.33] |
|  | CD127 mRNA | Transcriptomic | CNRQ | qRT-PCR | [0.39 - 1.07] |
|  | PD1 mRNA | Transcriptomic | CNRQ | qRT-PCR | [0.07 - 0.77] |
|  | IFNγ release after SEB stimulation | Functional test | pg/mL | ELISA | [99 – 4 959] |
|  | IL2 release after SEB stimulation | Functional test | pg/mL | ELISA | [119 – 2 872] |
|  | T cell proliferation after PHA stimulation | Functional test | % proliferating cells among T lymphocytes | FCM | [21 - 56] |
|  | T cell proliferation after OKT3 stimulation | Functional test | % proliferating cells among T lymphocytes | FCM | [4 - 43] |

**e-Table 2.**
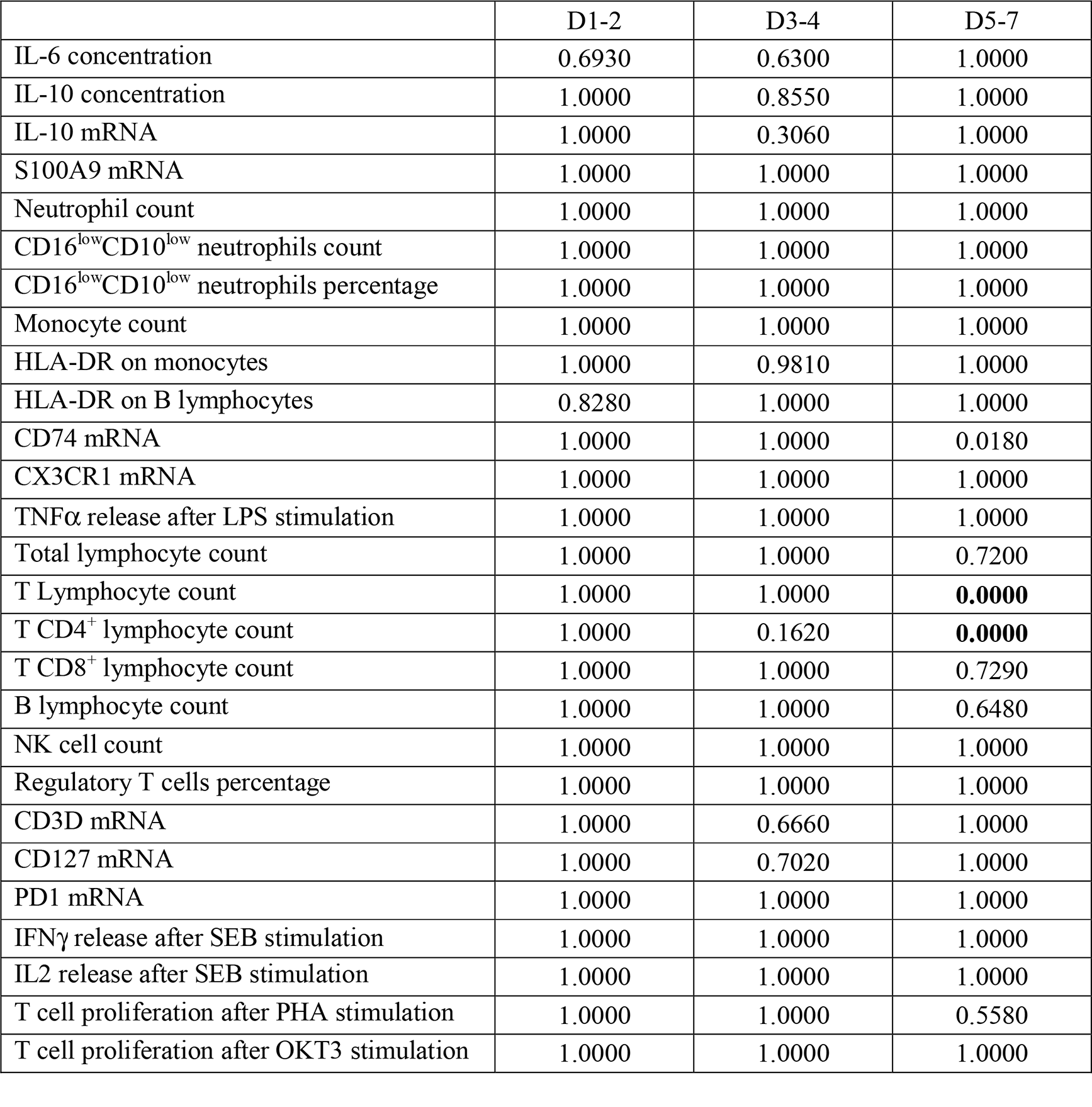
Analysis of covariance to evaluate the effect of group on immune response. Analysis of covariance (ANCOVA) was performed at each time point to characterize differences of marker level between each subgroup of critically ill patients. As some immune parameters were correlated to initial severity, we adjusted this analysis on age, SOFA score at admission and Charlson score and p-values were corrected for multi-testing using the Bonferroni method. The level of significance was set at 1%.

**e-Table 3.** Association between immune parameters and secondary infections at D30: univariate analyses. Three hundred and fifty three critically ill patients were prospectively included. Seventy-four patients developed at least one secondary infection within 30 days (D) and 16 died. The associations between D30 secondary infection and immune parameters (with death as competitive risk) were performed by implementing univariate Fine and Gray models at D1-2. Due to the insufficient number of deaths at the following time points, Cox proportional hazards models were used at D3-4 and D5-7. To allow comparison between models, hazards ratios (HR) calculated for each immune parameter were normalized to an increment from first to third quartile. Significant associations (bold print) between immune parameters and secondary infections were considered if p values ≤0.05.

| Immune Parameters | D1-2 |  | D3-4 |  | D5-7 |  |
| --- | --- | --- | --- | --- | --- | --- |
|  | HR (95%CI) | p | HR (95%CI) | p | HR (95%CI) | p |
| IL6 concentration | 1.00 (0.99-1.01) | 0.62 | 0.99 (0.92-1.07) | 0.844 | 1.00 (0.95-1.05) | 0.883 |
| **IL10 concentration** | **1.01 (1.00-1.01)** | **<0.001** | 1.04 (0.98-1.11) | 0.18 | **1.08 (1.00-1.17)** | **0.047** |
| IL10 mRNA | 0.94 (0.75-1.17) | 0.56 | 1.02 (0.83-1.26) | 0.846 | 1.12 (0.86-1.47) | 0.409 |
| **S100A9 mRNA** | 1.15 (0.91-1.46) | 0.23 | **1.57 (1.17-2.10)** | **0.002** | **1.59 (1.28-1.98)** | **<0.001** |
| **Number of neutrophils** | 1.12 (0.93-1.34) | 0.24 | 0.98 (0.73-1.31) | 0.885 | **1.37 (1.09-1.72)** | **0.007** |
| **Number CD16<sup>low</sup>CD10<sup>low</sup> neutrophils** | 1.23 (1.00-1.52) | 0.052 | 1.08 (0.85-1.36) | 0.531 | **1.18 (1.05-1.32)** | **0.005** |
| **% CD16<sup>low</sup>CD10<sup>low</sup> neutrophils** | **1.57 (1.03-2.40)** | **0.034** | 1.28 (0.93-1.76) | 0.132 | **1.35 (1.07-1.71)** | **0.011** |
| **Number of monocytes** | 1.16 (0.95-1.42) | 0.13 | 1.01 (0.74-1.40) | 0.933 | **1.29 (1.02-1.64)** | **0.034** |
| **Monocyte HLA-DR** | 0.90 (0.66-1.24) | 0.52 | 0.72 (0.50-1.03) | 0.069 | **0.35 (0.21-0.58)** | **<0.001** |
| **B lymphocyte HLA-DR** | 1.02 (0.72-1.44) | 0.9 | **0.69 (0.48-0.99)** | **0.043** | **0.58 (0.38-0.88)** | **0.011** |
| **CD74 mRNA** | 0.89 (0.66-1.20) | 0.45 | **0.58 (0.39-0.86)** | **0.006** | **0.45 (0.29-0.70)** | **<0.001** |
| **CX3CR1 mRNA** | 0.98 (0.74-1.31) | 0.91 | **0.62 (0.44-0.87)** | **0.006** | **0.62 (0.43-0.89)** | **0.009** |
| **TNFα after LPS stimulation** | 0.85 (0.60-1.20) | 0.36 | **0.66 (0.46-0.96)** | **0.03** | **0.49 (0.31-0.77)** | **0.002** |
| Number of lymphocytes | 0.94 (0.71-1.26) | 0.7 | 1.01 (0.73-1.41) | 0.947 | 0.95 (0.69-1.32) | 0.772 |
| Number of T lymphocytes | 1.00 (0.70-1.41) | 0.99 | 0.74 (0.52-1.05) | 0.097 | 0.86 (0.61-1.23) | 0.416 |
| Number of CD4+ lymphocytes | 0.95 (0.71-1.28) | 0.74 | 0.79 (0.55-1.13) | 0.188 | 0.86 (0.61-1.22) | 0.405 |
| Number of CD8+ lymphocytes | 1.10 (0.85-1.42) | 0.46 | 0.81 (0.60-1.09) | 0.167 | 0.94 (0.69-1.27) | 0.666 |
| Number of B lymphocytes | 1.11 (0.89-1.38) | 0.37 | 1.02 (0.80-1.31) | 0.857 | 1.06 (0.93-1.20) | 0.362 |
| **Number of NK cells** | **1.21 (1.01-1.46)** | **0.038** | 1.13 (0.93-1.37) | 0.21 | 1.11 (0.84-1.47) | 0.449 |
| % regulatory T cells | 1.05 (0.84-1.31) | 0.68 | 1.08 (0.84-1.38) | 0.552 | 0.98 (0.74-1.29) | 0.877 |
| **CD3D mRNA** | 0.92 (0.66-1.27) | 0.6 | 0.81 (0.56-1.17) | 0.265 | **0.50 (0.33-0.75)** | **0.001** |
| **CD127m mRNA** | 1.08 (0.84-1.40) | 0.54 | 0.86 (0.62-1.21) | 0.391 | **0.60 (0.38-0.93)** | **0.023** |
| **PD1 mRNA** | **0.64 (0.45-0.89)** | **0.009** | 0.93 (0.70-1.23) | 0.605 | 0.71 (0.48-1.03) | 0.071 |
| IFNγ after SEB stimulation | 0.79 (0.54-1.17) | 0.24 | 0.80 (0.60-1.07) | 0.137 | 0.68 (0.45-1.01) | 0.059 |
| IL-2 after SEB stimulation | 0.85 (0.64-1.15) | 0.29 | 0.92 (0.70-1.22) | 0.583 | 0.74 (0.51-1.06) | 0.101 |
| % T cell proliferation (PHA) | 1.13 (0.78-1.62) | 0.52 | 0.88 (0.65-1.19) | 0.399 | 0.93 (0.61-1.41) | 0.721 |
| % T cell proliferation (OKT3) | 1.05 (0.76-1.44) | 0.78 | 0.87 (0.62-1.20) | 0.394 | 1.07 (0.73-1.58) | 0.733 |

